# A database of pediatric drug effects to evaluate ontogenic mechanisms from child growth and development

**DOI:** 10.1101/2021.07.15.21260602

**Authors:** Nicholas P. Giangreco, Nicholas P. Tatonetti

**Affiliations:** Departments of Systems Biology and Biomedical Informatics, Columbia University, 622 W. 168^th^ Street, New York, NY, 10032

**Keywords:** Child Development, Data Mining, Pharmacology, Precision Medicine, Adverse drug events

## Abstract

Adverse drugs effects (ADEs) in children are common and may result in disability and death. However, current pediatric drug safety methods have not gone beyond event surveillance to identify and evaluate potential biological mechanisms. Children undergo an evolutionarily conserved and physiologically dynamic process of growth and maturation that can alter pharmacokinetics and pharmacodynamics. Our hypothesis is that temporal patterns of drug event reporting are reflective of dynamic mechanisms from child growth and development. We generated a database of 460,837 pediatric ADEs using generalized additive models (GAMs) that we have previously shown identify dynamic risk estimates of adverse drug events^1^. We identified 19,438 significant drug-event risks where drug risks corresponded with physiological development throughout childhood. Our results identified known pediatric drug effects and risk dynamics across child development that were not known previously. For example, we identified significant risk dynamics of montelukast-induced psychiatric disorders, including enriched risk (Odds Ratio 8.77 [2.51, 46.94]) within the second year of life. We developed a data-driven time-series clustering approach resulting in up to 95.2% precision and 97.8% sensitivity for categorizing risk dynamics across development stages for all ADEs including known but previously development-agnostic pediatric drug effects. We found that our real-world evidence may contain biologically-relevant underpinnings as well, where risk dynamics of CYP enzyme substrates were dependent on the enzyme’s expression across childhood. We curated this database for the research community to enable, for the first time, evaluation of real-world hypotheses of adverse drug effects across child growth and development.

## Introduction

Growth and maturation during child development are reflected by changes in the molecular landscape that may alter pharmacokinetics and pharmacodynamics^2^. In particular, cytochrome P450 enzymes, which metabolize 70-80% of drugs, exhibit dynamic activity during this time and within the first weeks of life these enzymes can vary up to 100-fold^3^. Activating metabolizers such as monooxygenases, aldehyde dehydrogenases, and amidases exhibit dynamic changes during infancy and early childhood^4^. During puberty, the hypothalamic-pituitary-gonadal axis orchestrates orders of magnitude increases of sex hormones like precursor estradiol^5, 6^. These hormonal dynamics drastically reduce or accelerate the bioavailability of drugs^7^ and regulate receptor availability and downstream signaling^6, 8^. Variations in metabolizing enzymes and other pharmacogenes will modulate drug and metabolite activities and increase the risk of adverse effects such as serotonin syndrome^9^ and NSAID-induced hypersensitivity^10^. Pharmacodynamic interactions can also lead to unexpected adverse effects, like paradoxical seizures following benzodiazepine treatment^11^ and heightened cyclosporin immunosuppression^12, 13^.

Despite strong evidence for molecular dynamics across child development and notable drug safety examples, such as doxorubicin-induced cardiotoxicity^14^ and methylphenidate-induced mental disorders^15^, the role of child development in pediatric drug effects remains largely a mystery. Preclinically, it is unclear if toxicity studies assess effects on child growth and development and significantly influence the design of trials involving children^16^. When pediatric trials are conducted (recently incentivized or required by regulation^17^), the studies include few patients in specific age groups within too short of a time period to sufficiently evaluate drug effects during child development^18^. This landscape is enabled by current drug safety legislation: drug safety can be extrapolated from adults to children if 1) the disease is determined to be similar and 2) the effect of the drug is similar between adults and children^19^. On the contrary, children are not simply small adults^20–22^ but can have distinct disease pathogenesis and trajectory^23^. Rapid growth and development during the period from birth through the teenage years complicates drug treatment when disease manifests. The pediatric drug safety landscape requires a large-scale approach, including the entire pediatric population, to ascertain the developmental contexts for this diverse population.

Real-world observational data, such as from spontaneous reports and electronic health record databases, can identify a diverse range of drug effects in large pediatric populations across different growth and development contexts^24^. Importantly, observational studies can identify idiosyncratic but clinically-relevant risks of medications taken throughout childhood^2^.

Notwithstanding the need to mitigate substantial bias inherent in real-world pediatric data, statistical and machine learning approaches can address this challenge and investigate the interaction between prescribed medications, observed side effects, and the developmental context of children. For newborns to young adults, and for those with diseases both common and rare, real-world observational data presents an opportunity to systematically investigate drug safety in the context of child development.

Current real-world observational data approaches in pediatric drug safety treat child development as independent periods of time instead of a continuous trajectory throughout childhood. The proportional reporting ratio (PRR), a disproportionality statistic of event prevalence associated with drug exposure, is commonly used by researchers and agencies to identify adverse drug events in the pediatric population. However, disproportionality statistics are limited to either modeling all children under 18 as one homologous group or to modeling risks within age groups which limits their power and treats children as distinct and unrelated to their similarly developing peers^25^. We have previously shown that generalized additive models (GAMs) address these limitations and generate robust and sensitive scores for modeling temporal adverse drug event risk across childhood^1^. GAMs allow for sharing information between development stages to reveal drug effect dynamics even when there may be scant evidence at a particular stage^1^. Moreover, GAMs are computationally efficient enough to apply in a high throughput manner to identify pediatric drug safety signals and evaluate the biology that may be associated.

We developed a resource of nearly half a million adverse drug effect risks across child development stages. We applied logistic generalized additive models (GAMs) to all observed pediatric adverse drug events (ADEs) in the FDA’s adverse event reporting system, generating drug effect risk estimates and dynamics across the stages of child development. Using GAMs, we mitigated reporting bias through covariate adjustment and increased more than two-fold detection of rare adverse events. Our ADE resource reproduced population-level physiological growth and development, and identified 19,438 significant as well as known, pediatric-specific adverse drug events. Our results identified known pediatric drug effects such as montelukast- induced psychiatric disorders, including enriched risk (Odds Ratio 8.77 [2.51, 46.94]) within the second year of life. A data-driven time-series clustering approach resulting in up to 95.2% precision and 97.8% sensitivity for categorizing risk dynamics across development stages for all ADEs including known but previously developmentally-unknown pediatric drug effects.

Furthermore, we evaluated and found evidence for observed drug risks associated with expression of cytochrome P450 enzymes across child development stages. We provide a resource for the pediatric drug safety community to further evaluate clinical and molecular hypotheses for observed ADE risks in the context of child development.

## Results

### Pediatric FAERS adverse drug event reporting

There were 264,453 pediatric reports in FAERS ranging from term neonates through late adolescents in the Pediatric FAERS dataset (Table 1). There were 460,837 unique drug-event (ADE) pairs reported over three decades. Majority of reports listed Female sex (52.9%). The most frequently reported drugs were from nervous system (35.3%), antineoplastic (26.8%), and alimentary tract and metabolic (13.5%) pharmacological classes. More than two drugs were reported for any given report (2.28 on average) and 95% of reports listed up to 8 drugs.

**Table 1:**
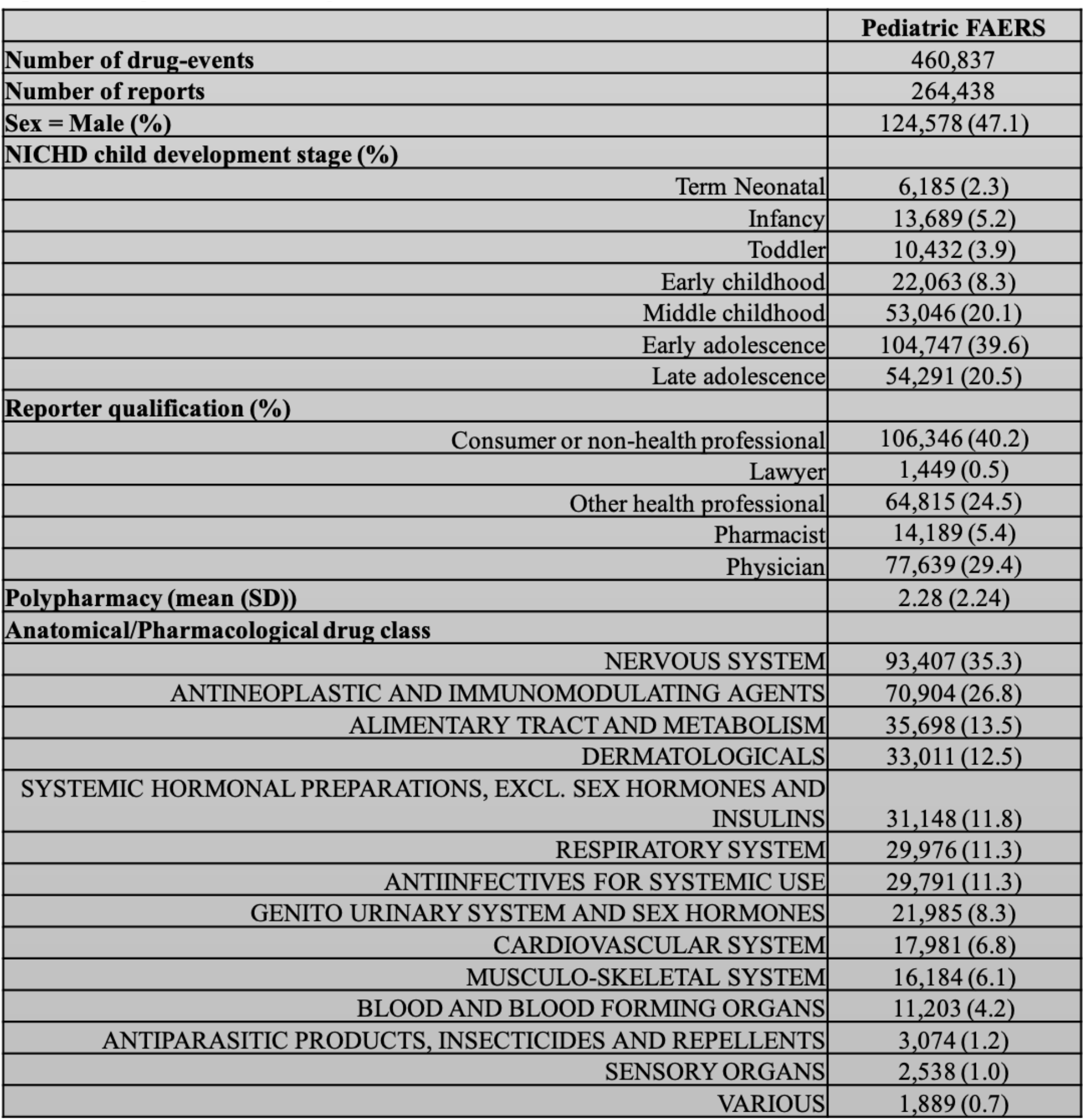
Adverse drug events across child development stages in Pediatric FAERS. The number and proportion of reports with the subject or drug characteristics. The drug class of the reported drug is in descending order.

### Our machine learning approach mitigates ADE confounding and reporting bias

In Pediatric FAERS, we observed 94% of ADEs were reported only up to 10 times in total (Figure S1A) and reporting factors such as stage, sex, reporting date, type of reporter, and class of drugs varied across childhood (Figure SB-F).

We evaluated GAM likelihood and generalizability by accounting for different factors or reporting characteristics for drug-events (see Methods). Compared to a base model, without covariates, accounting for sex across childhood resulted in a better model fit, specifically, a 2.1% and 16.9% increased model likelihood and generalizability, respectively (Figure 1A).

**Figure 1:**
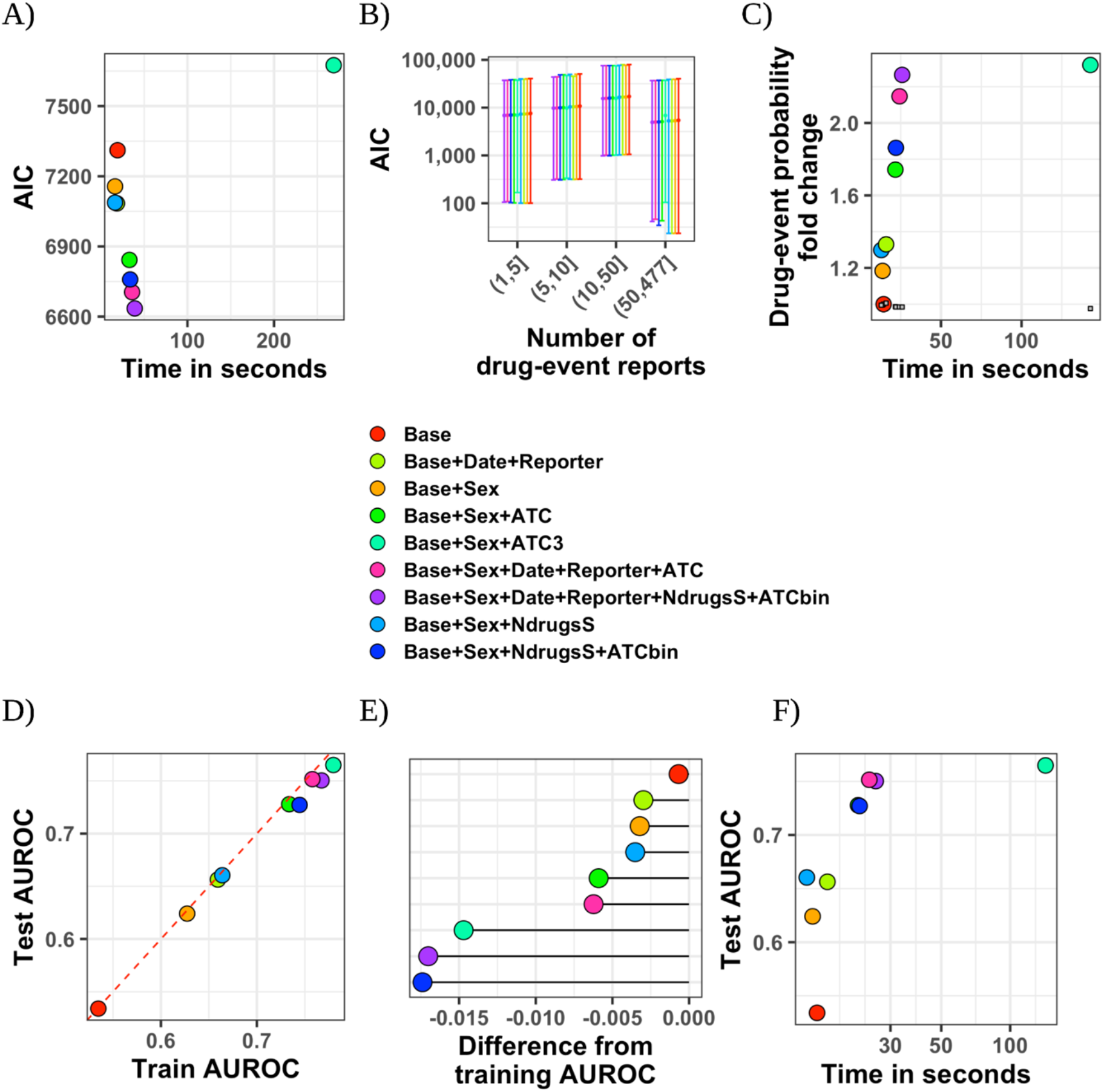
Generalized additive models (GAMs) generate robust and generalizable evidence for adverse drug events across childhood. A) The average Akaike’s Information Criterion (AIC) versus average time, in seconds, across drug-event GAMs, per model type. B) The AIC distribution of drug-event GAMs, per model type, between low to max reporting in drug-event sample. C) The drug-event probability average fold change from the ‘Base’ model. The control probabilities or drug-nonevent probabilities are shown in small squares for comparison. D) The average training versus testing area under the receiver operating characteristic (AUROC) curve for each model type. E) The average difference of testing from training performance or overfitting for each model type. F) The average testing AUROC versus the average time, in seconds, to fit the training data per model type.

Alternatively, accounting for the report date and type of reporter showed a 3.1% and 22.9% increased model likelihood and generalizability, respectively. The likelihood of the GAMs were stable in rank when considering different numbers of drug-event reports (Figure 1B). We found that separating the number of drugs taken (‘NdrugsS’) from their drug class (‘ATCbin’) slightly improved model likelihood (7.56% vs. 6.41% percent increase) and drug-event probability (86.3% vs. 74.3% percent increase) compared to integrating these two factors into one composite (‘ATC’) (Figure 1C). However, increasing the model complexity through separating rather than integrating factors showed an increase in model overfitting, defined as the training from testing AUROC difference, from -0.0062 to -0.017 (73% increase) while resulting in similar testing performance (Figure 1D,E). Overfitting was also exacerbated when increasing the number of drug classes (‘ATC3’). From these model comparisons, accounting for sex, report date, reporter, and the number of drugs within pharmacological classes produced the most improved model likelihood, generalization performance, and drug-event probability in a more modest time frame (Figure 1F).

### A resource of child development-specific ADEs

We generated 460,837 drug-event risk estimates across childhood for 10,770 and 1,088 unique adverse events and drug exposures, respectively. In comparison to the commonly used proportional reporting ratio (PRR), the GAMs estimated risk by sharing information across all stages (Figure 2A) that generated smooth risk relationships (Figure 2B). We then summarized the drug-event GAM risk distribution of drug-events across child development stages (Figure 2C). The model identified 152,919 or 33.2% drug-events that generated at least one nominally significant risk (GAM 90% lower bound beta coefficient>0) across child development stages (Figure 2D). To mitigate spurious associations, we further defined drug-event significance by comparing risk scores to a null risk from random drug and event associations (see Methods).

**Figure 2:**
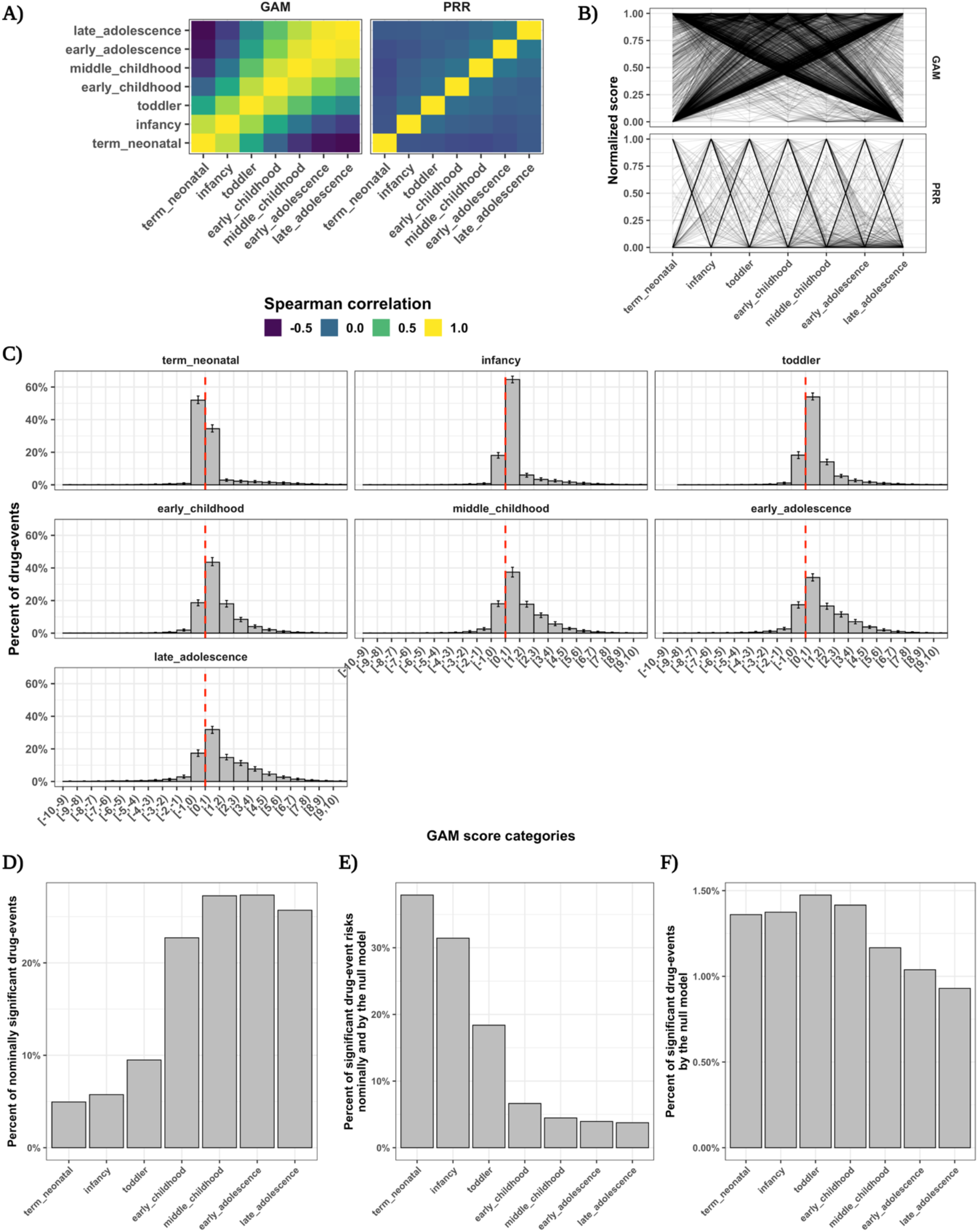
Drug-event GAMs estimate dynamic risks that share information across stages to identify pediatric adverse drug events. A) The spearman correlation between drug-event risks at child development stages from a random set of 2,000 drug-events, compared between our drug-event GAM and the popular proportional reporting ratio. B) The normalized risk scores across child development stages (normalized between [0,1] for each drug-event) from a random set of 2,000 drug-events, compared between our drug-event GAM and the popular proportional reporting ratio. C) The percent of nominally significant drug-event risks (90% lower bound above 0 or the null association) across child development stages for the drug-event GAMs. Error bars represent the 95% confidence interval for percentages calculated across 100 bootstraps of drug-events from Pediatric FAERS. The dashed redline indicates the null association between an adverse event and drug exposure across child development stages. D) The percent of nominally significant drug-event risks (90% lower bound above 0 or the null association) across child development stages. E) The percent of significant drug-event risks by the null model, out of all nominally significant drug-event risks, across child development stages. F) The percent of significant drug-events by the null model out of all drug-events in Pediatric FAERS.

This narrowed our findings to 19,438 or 4.2% of drug-events passing the 99^th^ percentile null risk threshold at each stage. The percent of risks exceeding both nominal and null model significance thresholds were higher for earlier child development stages (Figure 2E). We defined significant drug-events where at least one risk passed nominal and null model significance thresholds (Figure 2F). We found significance by the null model detected two times more drug risks for pediatric adverse events compared to nominal significance (odds ratio 2.00 [1.85, 2.15]).

### Drug-event GAMs identify known pediatric drug effects

We evaluated drug-event risk scores within known, or positive, drug-events compared to unassociated, or negative, drug-events. We first examined risks across childhood between 312 positive and negative drug-events, which were not specific to the pediatric population, in the Ryan et al. reference set. There was a 1.15-fold higher rate (relative risk 90% CI [0.72, 1.84]) of nominally significant risks for positive compared to negative drug-events. We then examined risks across childhood for 187 positive and negative drug-events, which were specific to the pediatric population, curated by the Global Research in Pediatrics (GRiP) consortium. There was a 1.9-fold higher rate (relative risk 90% CI [1.08, 3.38]) of nominally significant risks at child development stages for positive compared to negative drug-events. We determined drug-event risk at stages, which were unspecified previously, and found 22 (29.3%) drug-events contained nominally significant risk scores and displayed dynamic drug-event risk across childhood (Figure 3).

**Figure 3:**
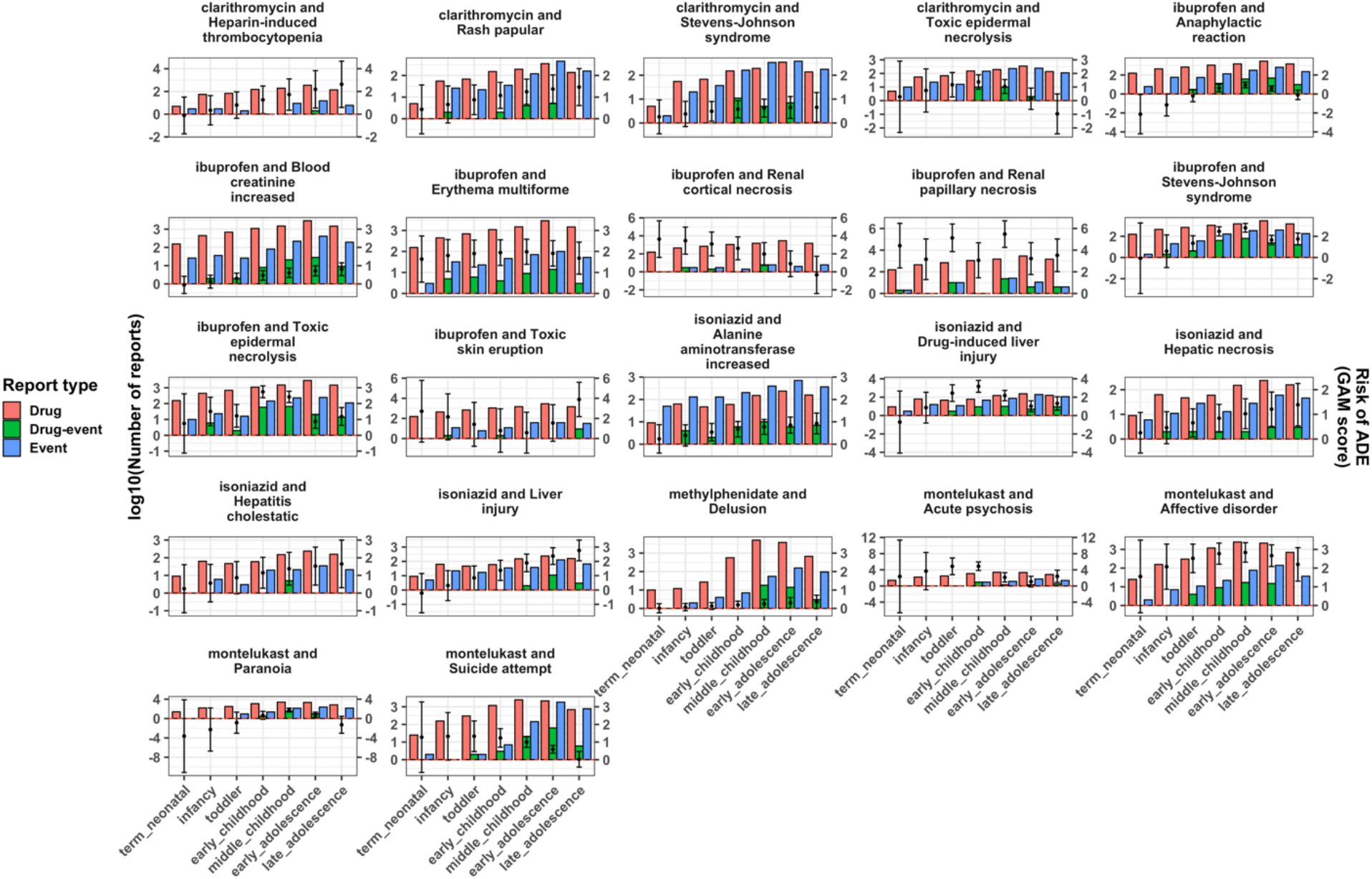
Known pediatric drug effects show dynamic risk across child development stages. Adverse event risk across child development stages for drug-events in the GRiP drug-event reference set with either epidemiological or mechanistic evidence in children.

### Drug effects show dynamic risks by effect etiologies and drug mechanisms

We evaluated the enrichment of 1,518 medication and drug classes showing significant risks for 8,675 MedDRA adverse events and disorders at child development stages (Figure 4A). We found 282 or 0.13% of drug effects were enriched (FDR<0.05) for significant drug-events at child development stages (Figure 4B). Adverse events and systemic disorders were found for both medications and drug classes (Figure 4C). Of note, we identified 32 medications significantly associated within systemic disorders (MedDRA system organ class) at child development stages (Figure 4D and Table S1).

**Figure 4:**
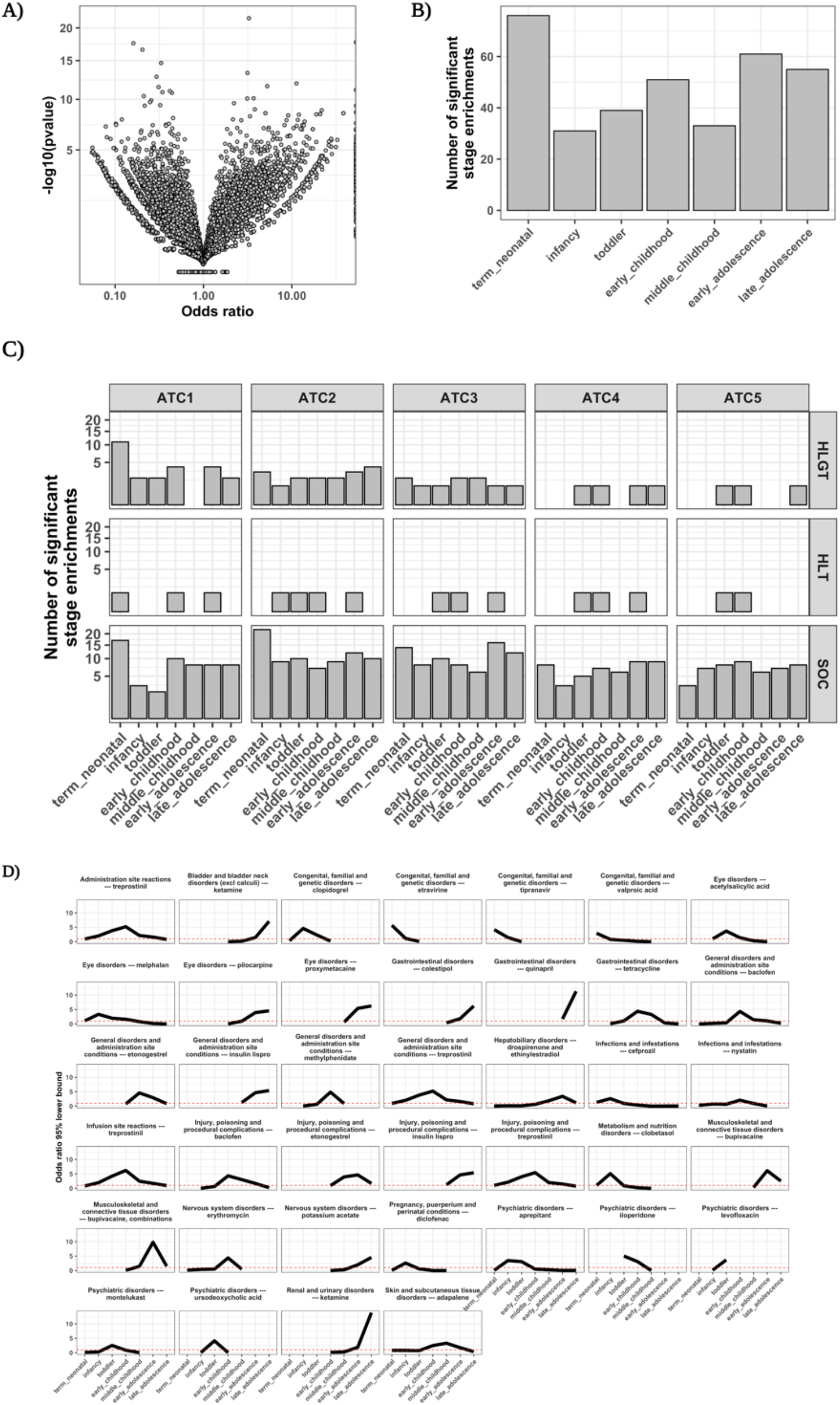
Drug risks and adverse effects are associated to child development stages. A) Volcano plot of the enrichment for drug-events among ATC and MedDRA classes within child development stages. B) The number of significant enrichments at each stage (false discovery rate of 0.05) for significant drug-events. C) The number of significant enrichments by the ATC and MedDRA class. D) The 95% lower-bounded odds for disorders by medications with at least one significant enrichment across child development stages. The presence of the line indicates significant drug-events were present at that stage, and none were significant otherwise. The red dashed line indicates the null enrichment threshold. Abbreviations: ATC: Anatomical Therapeutic Class; ATC1-5: ATC 1^st^ level – 5^th^ level; SOC: System Organ Class, HLGT: Higher-Level Group Term, HLT: Higher-Level Term.

Figure S2 shows the percentage of significant drug events and their odds enrichment within each development stage for a drug effect etiology. Younger children were at a greater risk (odds ratio 3.19 [2.83, 3.58]) for experiencing congenital disorders, where about 60% of congenital-related drug-events were significant during the first month of life compared to other development stages. Infants were at a greater risk (odds ratio 1.45 [1.20, 1.74]) of experiencing drug effects related to surgeries and medical procedures. We found that nervous system disorders (odds ratio 1.16 [1.06, 1.27]) and eye disorders (odds ratio 125 [1.09, 1.43]) became significant risks just after the second year of life. Endocrine disorders became significant drug effect risks (odds ratio range 1.36 - 1.58) during the stages of early childhood through early adolescence which parallels the biological processes of puberty. Only during late adolescence were gastrointestinal disorders significant drug effects (odds ratio 1.24 [1.10, 1.40]).

Figure S3 shows the percentage of significant drug events and their odds enrichment within each development stage dependent for drug pharmacological class. Younger children were at significant risk for drug effects from cardiovascular drugs (odds ratio 1.29 [1.16, 1.43]), mirroring the significant risk for cardiac disorder effects (odds ratio 1.57 [1.39, 1.79]).

Significant drug effect risk from antineoplastic agents were significantly enriched in each development stage after the first month of life (odds ratio range 1.12 – 1.23). Moreover, mirroring hormonal fluctuations that occur during puberty, we found significant enrichment of drug risks from those affecting the genito-urinary system and sex hormones (odds ratios range 1.28-1.38).

### ADEs had dynamic risk patterns across child development

We found drug-event GAM risk followed distinct patterns across child development stages. However, while most risk patterns were found to be directly increasing (65%) or decreasing (28.6%), there were 28,892 (6.4%) drug-events that fell outside these trends and remain unassigned.

We performed a data-driven clustering strategy to assign patterns for drug-events in Pediatric FAERS. We evaluated our clustering strategy to categorize injected ‘canonical’ drug-event risk patterns, including increase, decrease, and plateau patterns, and be both sensitive and precise towards cluster assignments (see Methods). We found that clustering algorithms, using different combinations of distance and centroid parameters, assigned canonical patterns to clusters with up to 95.2% precision (cluster purity) and 97.8% sensitivity (dynamics localization; Figure 4A). Increasing the number of identified clusters, even with reduced sensitivity, increased the precision for canonical dynamics (Figure 4B). Moreover, treating the drug-event risks as time-series or shapes that share information across childhood resulted in overall higher clustering sensitivity and precision (Figure 4C).

We compared all pairwise drug-events in Pediatric FAERS using shaped-based distance and partitional clustering, and found the dynamics clustered into four visually distinct risk dynamics across childhood (Figure 4D). While 137,008 (65.25%) drug-events were assigned to the increase dynamic cluster, only 5,397 (1.79%) were significant by the null model. In contrast, 20,127 (4.37%) drug-events were assigned to the plateau dynamic cluster and 5,885 (29.23%) of those were significant by the null model (Figure 4E).

We visualized the cluster assignments per the 32 drug risks for systemic disorders enriched within child development stages. We identified dynamic risk patterns of known drug risks, such as montelukast-induced psychiatric disorders (Figure 4F). There were drugs that posed only one risk dynamic, such as quinapril exposure and gastrointestinal disorders, and drugs that posed multiple risk dynamics for specific sub-etiologies, such as valproic acid and congenital disorders (Figure S3).

### Cytochrome P450 expression dynamics were predictive of drug effect risks across child development stages

We took a systems biology approach to evaluate our real-world observations for putative biological mechanisms during growth and development. For example, dynamics of cytochrome P450 enzymes expression across child development stages may alter systemic effects of drugs we observe in Pediatric FAERS (Figure 6A). We found that grouping significant drug-events by the *CYP* metabolizing enzyme of the drug showed enrichment (fisher exact test FDR<0.05) within child development stages (Figure 6B). We then hypothesized the expression of *CYP* enzymes across child development stages is associated with observed dynamics of systemic risks by drug substrates.

**Figure 5:**
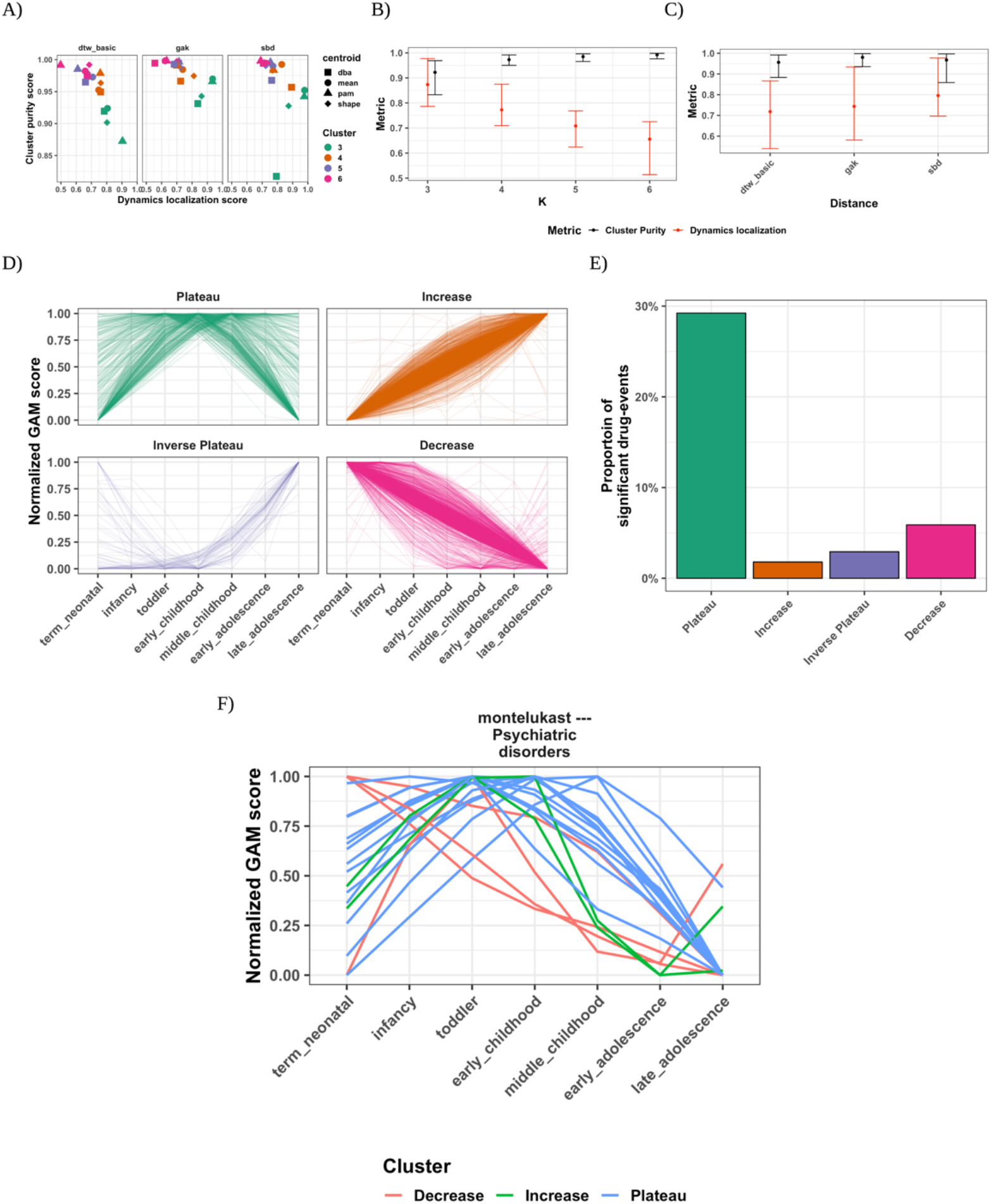
Time-series based clustering categorizes drug risks into dynamic patterns across child development. A) Clustering metrics dynamics localization versus cluster purity scores across clustering models for (Number of clusters, centroid, distance) triplet hyperparameter sets. B) Cluster metric scores and their 95% confidence interval versus the number of clusters to fit in the clustering model. C) Cluster metric scores and their 95% CI versus the drug-event distance used to fit in the clustering model. See Methods for details on the clustering strategy and metrics. D) Cluster assignments assigned to putative risk dynamics categories after fitting top cluster model with all drug-events in pediatric FAERS. The GAM coefficients were normalized between [0,1] producing scores across child development stages for each drug-event. E) Percent of drug- events assigned from the top cluster model that were significant by the null model. F) Montelukast-psychiatric disorder drug-events assigned risk dynamics clusters.

**Figure 6:**
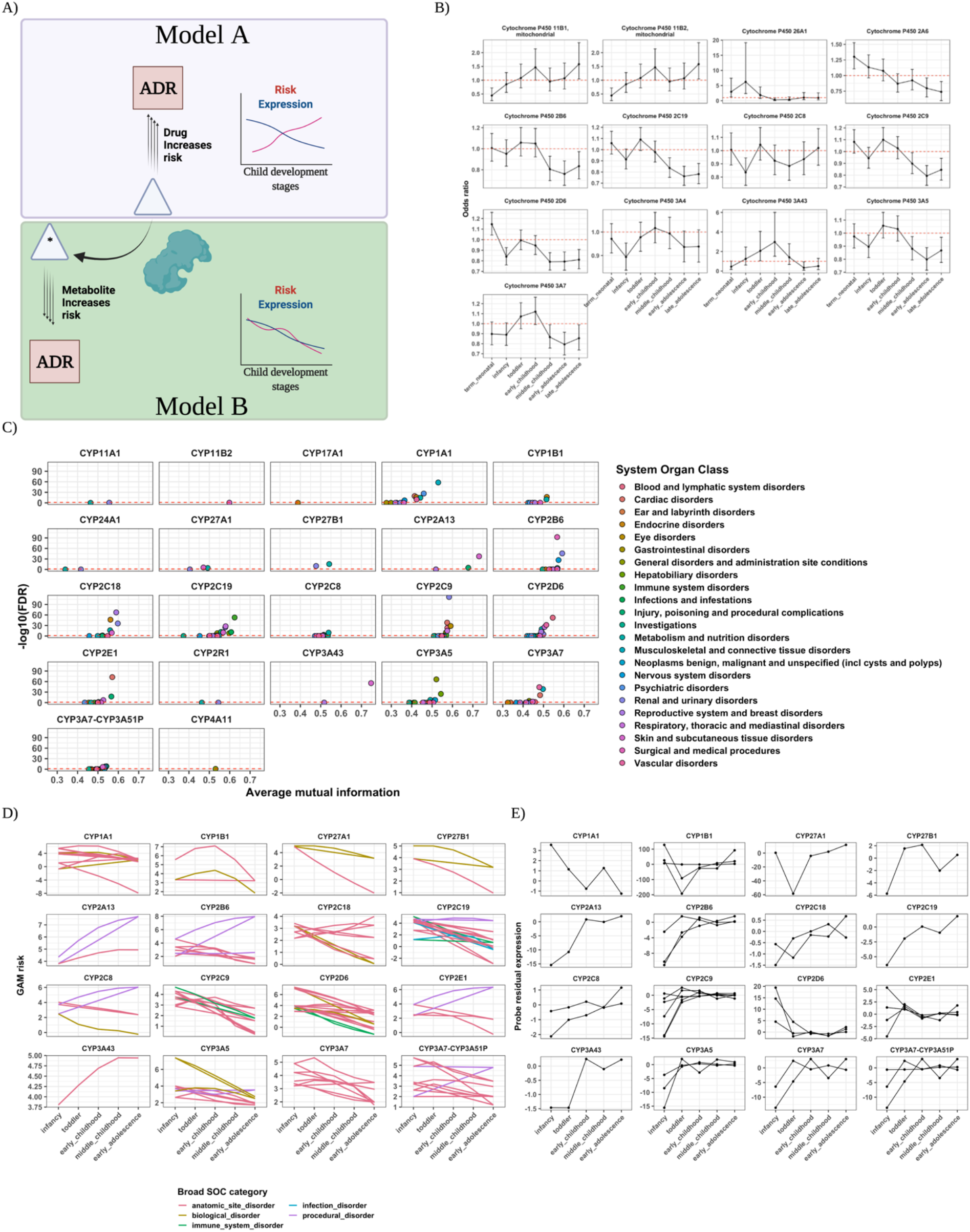
Drug risks from real-world observations associate with gene expression dynamics across childhood. A) Putative metabolic models of development-dependent risk. Adverse drug reactions (ADRs) can be influenced by dynamic metabolic processes during growth and development. In Model A, the risk of an ADR stems from increased concentration of the drug. The decreased metabolism, such as by the enzyme in green, increases the bioavailability of the drug resulting in the observed ADR. In this case, drug risks are inversely proportional to expression dynamics of this enzyme across child development stages. In Model B, the risk of an ADR stems from the aberrant modification of the drug such as by the enzyme in green. The concentration of the metabolite results in an observed ADR. Drug risks are directly proportional to expression dynamics of this enzyme across child development stages. B) Enrichment of significant drug-events at child development stages out of all drug-events with cytochrome P450- metabolized drugs. The red dashed line indicates the null enrichment threshold, and the error bars are the 95% confidence intervals of the odds ratio. Shown are CYP enzymes where at least one stage was significantly enriched for CYP-metabolized drug risks across child development stages. C) Volcano plots of the average mutual information (MI) for drug systemic risks by CYP substrates versus the -log10 t-test false discovery rate (FDR) between substrate and non-substrate mutual information. A dashed red line for FDR=0.05 is shown. D) The average drug substrate risk for systemic risks for each CYP enzyme sharing significant information (t-test FDR<0.01) within systemic disorders. E) The residual probe expression across child development stages for the CYP enzymes in B).

We first constructed a dataset of gene expression across childhood (Table S2 and Figure S5A,B) where previously identified age-associated genes by Stevens et al. were corroborated in our dataset following our re-normalization and adjustment procedure (Figure S5C and see Methods). Genome-wide expression did not significantly change on average between infancy through early adolescence stages (t-test FDR>0.05). We identified 22 *CYP* genes comprising 50 probes, with concordant probe expression within genes, that showed dynamics across child development stages.

We evaluated shared dynamic information between adverse drug event risks and *CYP* enzyme expression across child development stages. Specifically, we compared the mutual information between *CYP* gene expression and drug risks by *CYP* substrates compared to non-substrates, accounting for risk estimate variation, across child development stages. Out of 780 significant drug-events present on drug product labels, there were 429 drug-events where the drugs were substrates of the 22 *CYP* enzymes. We found dynamic expression of *CYP2C18* (t-test p-value<5.57E-23) and *CYP27B1* (p-value<1.69E-31) were more informative of drug risk dynamics by substrates compared to non-substrates across all side effects. However, we found enzyme expression significantly associated with (t-test FDR<0.05) drug substrate risks across multiple systemic disorders (Figure 5C). Specifically, we found systemic risks by drug substrates for 16 *CYP* enzymes contained significant shared information across child development stages (Figure 5D,E).

## Discussion

### Study overview

We systematically generated 460,837 observed adverse drug event risk estimates across child development stages. Unlike disproportionality measures that quantify risk within age groups, our approach using generalized additive models (GAMs) both mitigated bias and shared information to estimate risk across stages. We assessed the impact of different factor combinations and specified GAMs that produced robust and generalizable adverse risk estimates. This approach, for the first time, systematically identified adverse drug event risk dynamics across childhood in all stages of child development.

### Biologically-inspired modeling using drug-event GAMs

GAMs enable estimating dynamic risks by sharing information across childhood. GAMs are common in evaluating environmental and ecological characteristics over space and time, such as black smoke particulate exposures across the UK over four decades^26^ and artificial light density on bird stopover in different habitats during autumn migration^27^. The aim of pharmacovigilance activities, on the other hand, has been to detect signal for potential and rare adverse effects from marketed medicinal products^28^. Disproportionality methods have served this monitoring purpose, and for the pediatric population the common practice of stratifying data into age groups was shown to uncover signal that would otherwise go unnoticed^25^. Alternatively, our approach goes beyond monitoring and towards evaluating drug effects over time as children grow and develop on molecular, physiological, and structural levels. We flip-the-script in pediatric drug safety by developing and applying a strategy informed by pediatric developmental biology.

### Drug-event GAMs were more robust and predictive of pediatric adverse events

Our models quantify relationships for every drug and adverse event co-reported across pediatric age groups as opposed to isolated pediatric groups. In this way, we were able to extract dynamic risk information across child development stages instead of concatenating isolated risks. Despite including a quarter of a million reports with very few events co-reported with a drug in our models, we extracted useful information based on 1) increased model fit to the data when considering important factors that contribute to adverse drug events^29, 30^ and 2) increased generalizability for a hold-out test set with modest overfitting. This robust, systematic approach generated hypotheses of pediatric drug effects including rare adverse drug events.

### Drug-event GAMs identify known pediatric drug effects

We identified putative risk dynamics for known pediatric drug effects. We generate evidence of the risk for all pediatric-specific adverse drug events curated by the Global Research in Pediatrics (GRiP) consortium^31, 32^. We provide a temporal map of the risk for these pediatric drug-events across child development stages that was unspecified previously. For known culprits such as montelukast, our approach identified significant risk during mid-childhood (Figure 3) corresponding to studies from the Swedish ADR database^33, 34^ and the World Health Organization’s Vigibase^35^. Moreover, our unsupervised clustering approach further evaluates the dynamics of these and thousands of drug risks that were previously unknown. We categorize dynamics of drug-events that visually correspond to their temporal trend across child development stages, as expected. Importantly, our systematic approach generates evidence for classes of medications resulting in seemingly distinct but temporally-related side effects.

### Drug-event GAMs cluster within robust risk dynamic patterns across child development

We investigated patterns of half a million drug risks across all drugs and adverse events in the population. We optimized our clustering procedure to accurately and precisely cluster known (through simulation) drug risk dynamics. Our procedure rediscovered the canonical (through our simulation study^1^) cluster ‘inverse plateau’ that we intentionally left out, which signifies its ability to generate clusters of canonical trends over time. Cluster assignments can now be utilized to test hypotheses for enriched drug risk dynamics by common therapy, disease, and type of side effect. This can only be done by having large amounts of drug risks available. Our novel data mining methodology is the first approach to evaluate when during childhood side effect risk may be occurring for a particular drug therapy or drug class. From thousands of drug effects, we can investigate effects arising from medications with shared pharmacology.

### Drug-event GAMs facilitate evaluating ontogenic-mediated pediatric drug effects

Our database enables for the first time the ability to investigate pediatric, ontogenic biology from observational data. For example, the cytochrome P450 enzymes, which metabolize about 70-80% of drugs on the market, exhibit dynamic changes in activity across child development that result in altered drug actions and effects^9, 36^. Notably, these enzymes show characteristic activity patterns where it is generally thought *CYP* enzymes surge in activity during the first few years of life and then gradually decline to mature levels^37^. From our approach, we showed drug risks within stages throughout childhood are enriched when metabolized by *CYP* enzymes. We curated a gene expression dataset to evaluate a biological underpinning or shared information between our observed risks and the expression of drug enzymes across child development stages. We found evidence for ergocalciferol, also known as vitamin D2, resulting in risks for Polyuria, Hypercalciuria, and Hypervitaminosis D from *CYP27B1* metabolism, where ergocalciferol treatment was previously shown to result in abnormally high calcium levels in younger pediatric patients^38^. We also identified evidence of drug risk dynamics associated with *CYP2C18* expression, where the ontogeny of this gene isoform is not known but drug binding was found to be different compared to other *CYP2C* gene family isoforms^39^. There were four drug-event substrates different from other *CYP2C* family substrates, comprising the three drugs cyclophosphamide, ifosfamide, and omeprazole. The alkylating agents ifosfamide and its parent compound cyclophosphamide are known to share a toxicity profile of myelosuppression and urotoxicity^40^, but our data suggest and corroborate the long term effect of severe cellular damage and possible infertility in pediatric cancer-survivors^41^. Moreover, our observational analysis corroborates omeprazole risk for the benign adverse effect of enlarged breast tissue in neonates that stems from increased oestrogen production^42^ (Figure 7). Our resource allowed for generating clinically-relevant molecular and developmental hypotheses found through a systematic data mining approach.

**Figure 7:**
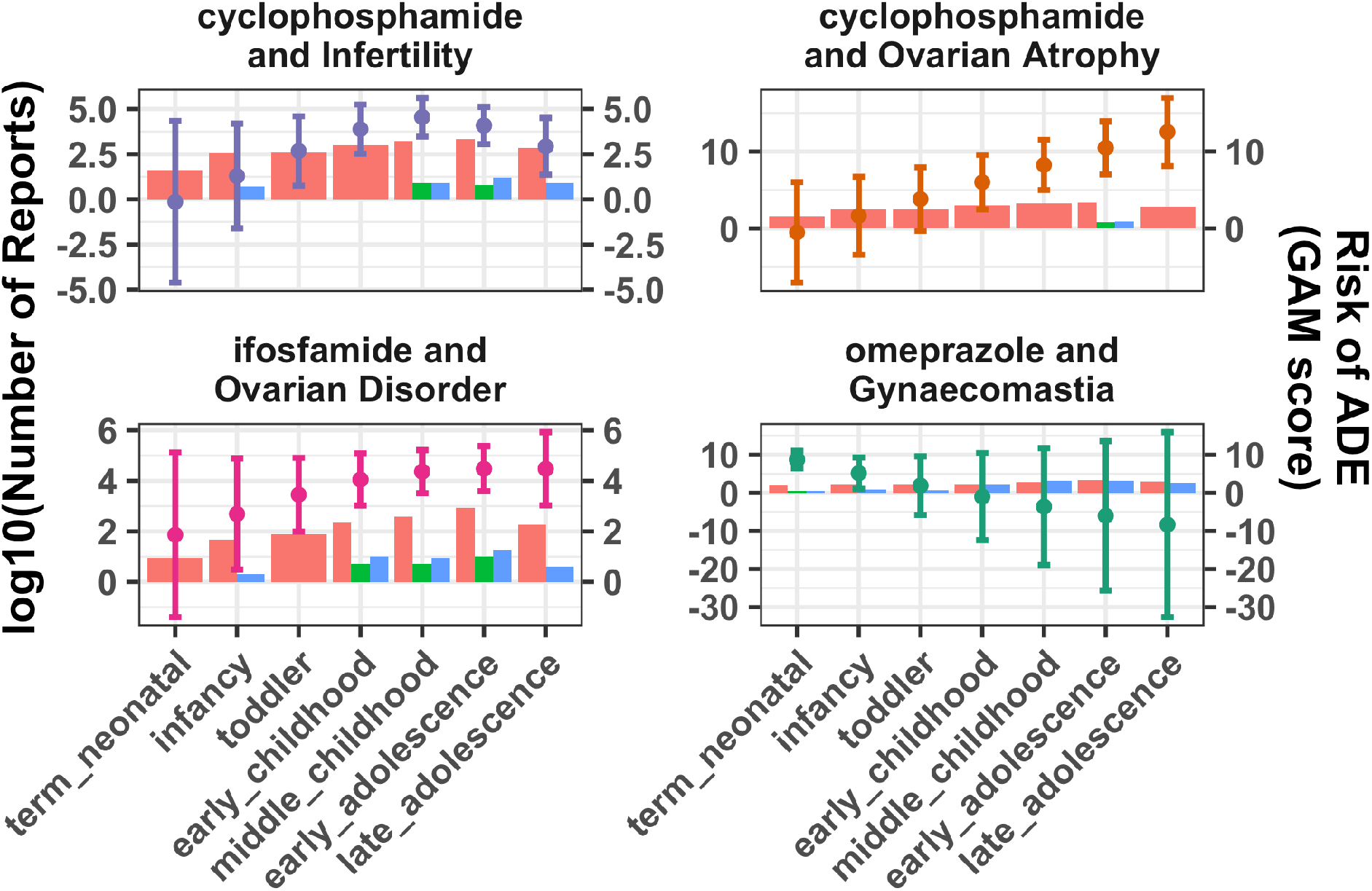
Adverse event risk dynamics by CYP2C18 drug substrates associate with the enzyme’s expression dynamics. We found CYP2C18 as one of the only two CYP enzymes significantly associated with adverse risks of their substrates. The drugs in each drug-event are metabolized only by CYP218 and no other CYP2C family gene. Each drug-event is reported in each facet with different colors. Bars are in red, blue, and green to indicate the number of drug, event, and drug-event reporting, respectively, in Pediatric FAERS. Related to Figure 6.

### Pediatric drug effect resource

We made available, for the first time, a database of half a million pediatric drug effects across growth and development stages. We have shown how a massive data mining effort captures known pediatric drug effects and reproduces system-wide development patterns. We included standardized drug and adverse event vocabulary information as well as derived clustering and stage enrichment data to further investigate both clinical and molecular pediatric ADE hypotheses. Importantly, we include a summary of the covariates within our significant drug-event GAMs to illuminate the data-context, as well as promote equitable treatment of our predictions, for which we generated these hypotheses. Our resource is novel in both its accessibility to the clinical community but also to researchers for evaluating mechanistic hypotheses such as metabolic associations on observed drug risk dynamics as shown here for substrates of *CYP* enzymes. We include additional data for evaluating shared information between drug risks and their targets, carriers, transporters, and enzymes. We make available our findings to the research community to accelerate pediatric drug safety research.

### Limitations

This study has some limitations. First, the observations of drug reporting over time in FAERS contains biases and confounding factors that may impact our risk estimates. While we mitigate potential biases by including confounding factors in our GAMs, we acknowledge that our models may still contain these biases. For example, we did not take into account the administration route of the drugs, which are known factors in drug toxicity. This treatment characteristic was rarely present in our data and so we could not adequately consider this factor in our drug-event GAMs. Though our model evaluation strategy was not exhaustive in performance metrics and model factors, we showed reduced bias and increased signal when accounting for known adverse drug event risk factors. We represented child growth and development through pre-defined stages with age ranges defined by NICHD which were intended to standardize consistent age groups for randomized clinical trials. These stages, however, remain temporally-related and were evaluated by multiple stakeholders such as the American Academy of Pediatrics and the Centers for Disease Control and Prevention. Our approach generated nonzero risk at stages for drug-events with no reporting. While it is difficult to interpret nonzero risk estimates at stages with no occurrence of a report, the sharing of information between stages including report and drug characteristics promoted risk estimation. Nevertheless, nonzero risk estimation from no explicit reporting at a stage may provide an opportunity to alleviate the burden of underreporting of medication side effects. While we performed a stringent strategy to generate *CYP* expression that corresponded with the original Stevens et al. findings, our latent representation was not validated against another data source. Nonetheless, this triangulation procedure augments real-world evidence to pinpoint putative metabolic mechanisms for our observed drug risks. Our resource not only lends itself to assess when during childhood risks may occur, but also toxicities arising from shared drug pharmacology furthering our understanding of adverse drug effect mechanisms and developmental pharmacology.

## Conclusion

Children may be prescribed medications at any point during childhood, and we provide the first resource to identify and evaluate drug effects across child development stages. We generated robust risk estimates for half a million adverse drug events by sharing information across child development stages. The generated risk estimates identified known pediatric drug effects apart from risk estimates from unknown drug and event associations. These risk estimates spanned across the child development stages to allow for examining differential drug-event risk across childhood, such as by systemic disorders and drug mechanisms. Moreover, our real-world evidence was shown to contain dynamic information on putative biological mechanisms that supports evaluating adverse drug effect hypotheses. We provide a multi-purpose resource for the community to explore from identifying safety endpoints in clinical trials to evaluating known and novel developmental pharmacology.

## Data Availability

https://github.com/ngiangre/pediatric_ade_database_study

## Acknowledgements

We thank the members of the Tatonetti lab and Dr. Jon Elias for their feedback. NPG and NPT are funded by R35GM131905. Figures were developed using Biorender.

## Authors’ contributions

NPG and NPT designed the study and performed analyses. NPT provided supervision and critical feedback on the manuscript. NPG wrote code. The authors read and approved the final manuscript.

## Figure legends

**Figure S1:**
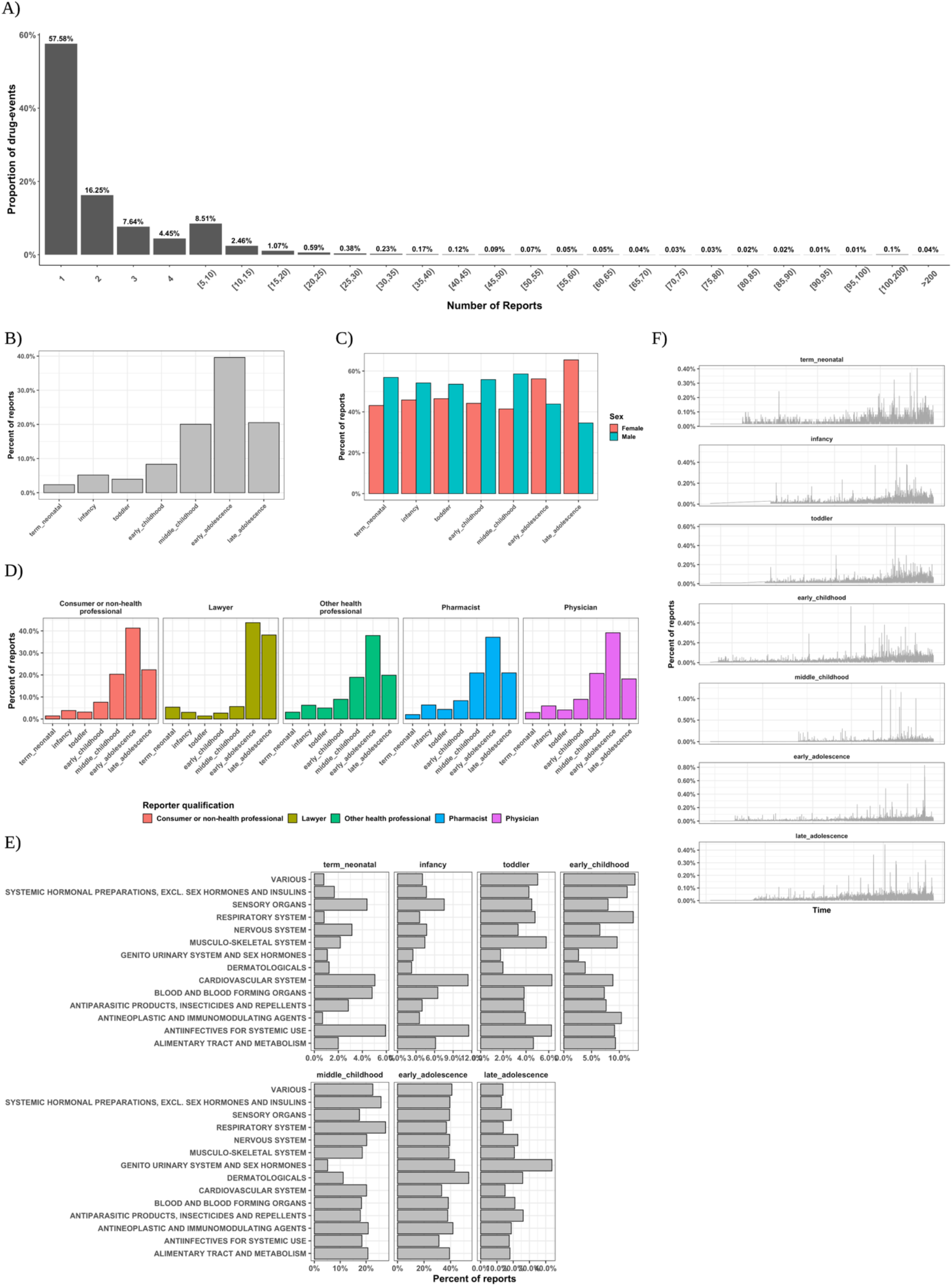
Observed adverse drug events vary in reporting characteristics across child development stages. A) The proportion of adverse drug events or drug-events out of all unique drug-events with a number of reports in pediatric FAERS. B) The percent of drug-events across child development stages by C) sex, D) drug-event reporter qualifications, E) drug class, and F) date, spanning between 1990 and 2020, of drug-event reporting. Related to Table 1.

**Figure S2:**
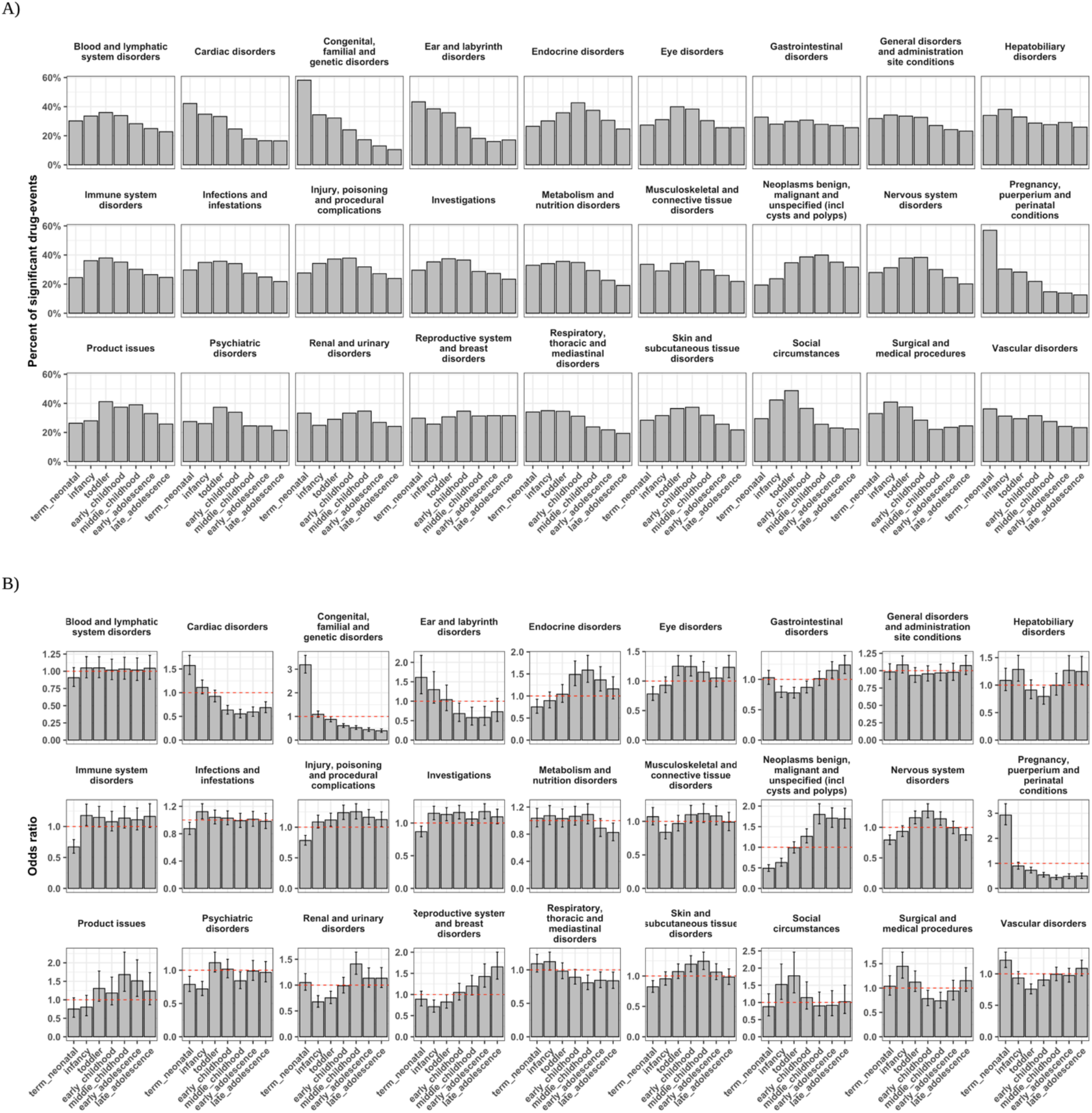
Drug risks vary by etiology across child development stages. A) The percent of significant drug-events, by the null model, at child development stages out of all drug-events within anatomical therapeutic classification (ATC) level 1 pharmacological classes. B) The enrichment of significant drug-events by pharmacological class at child development stages. The red dashed line indicates the null enrichment threshold. Related to Figures 2 and 4.

**Figure S3.**
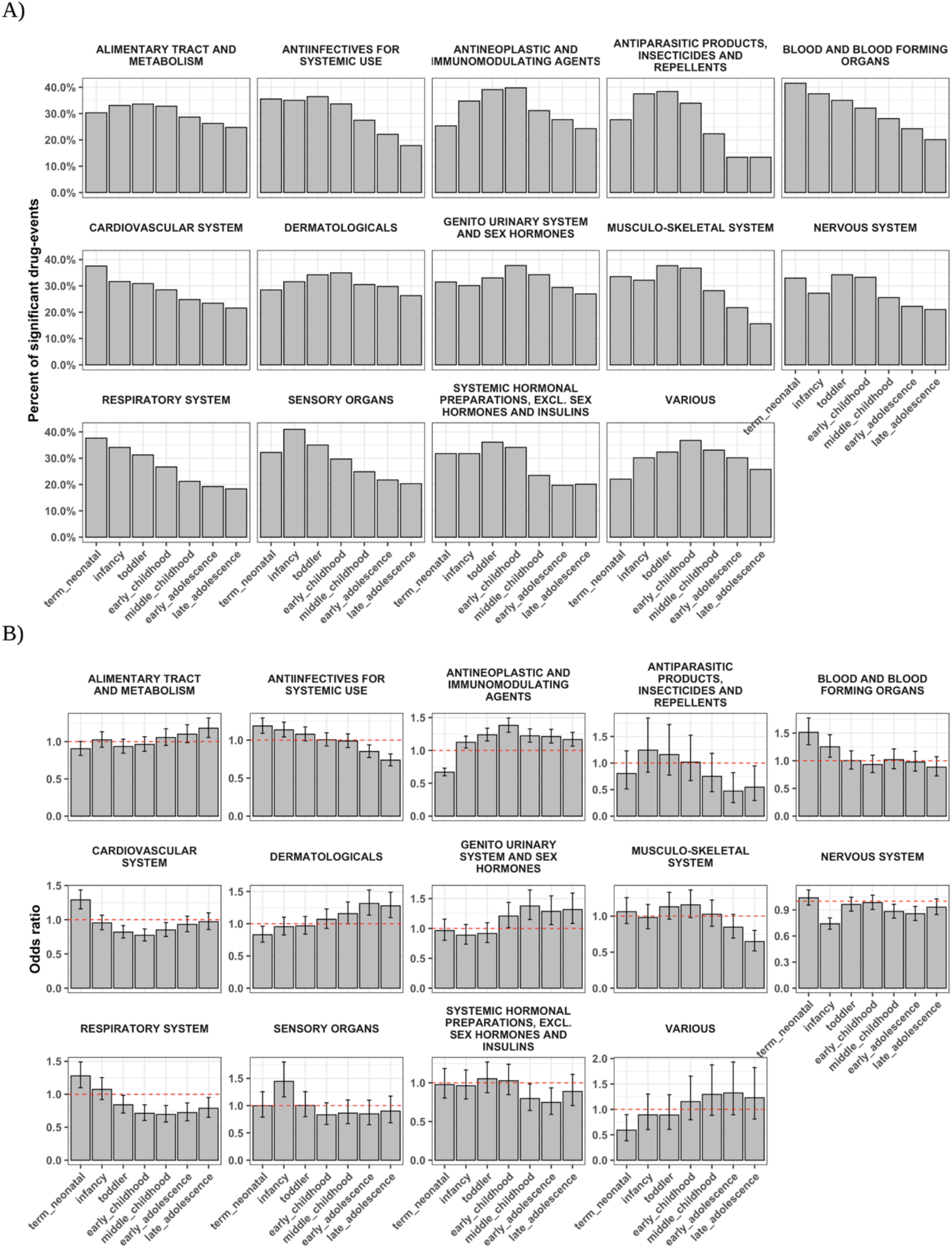
Drug risks vary by drug mechanisms across child development stages. A) The percent of significant drug-events, by the null model, at child development stages out of all drug- events within MedDRA system organ class (SOC) etiologies. B) The enrichment of significant drug events by SOCs at child development stages. The red dashed line indicates the null enrichment threshold. Related to Figures 2 and 4.

**Figure S4:**
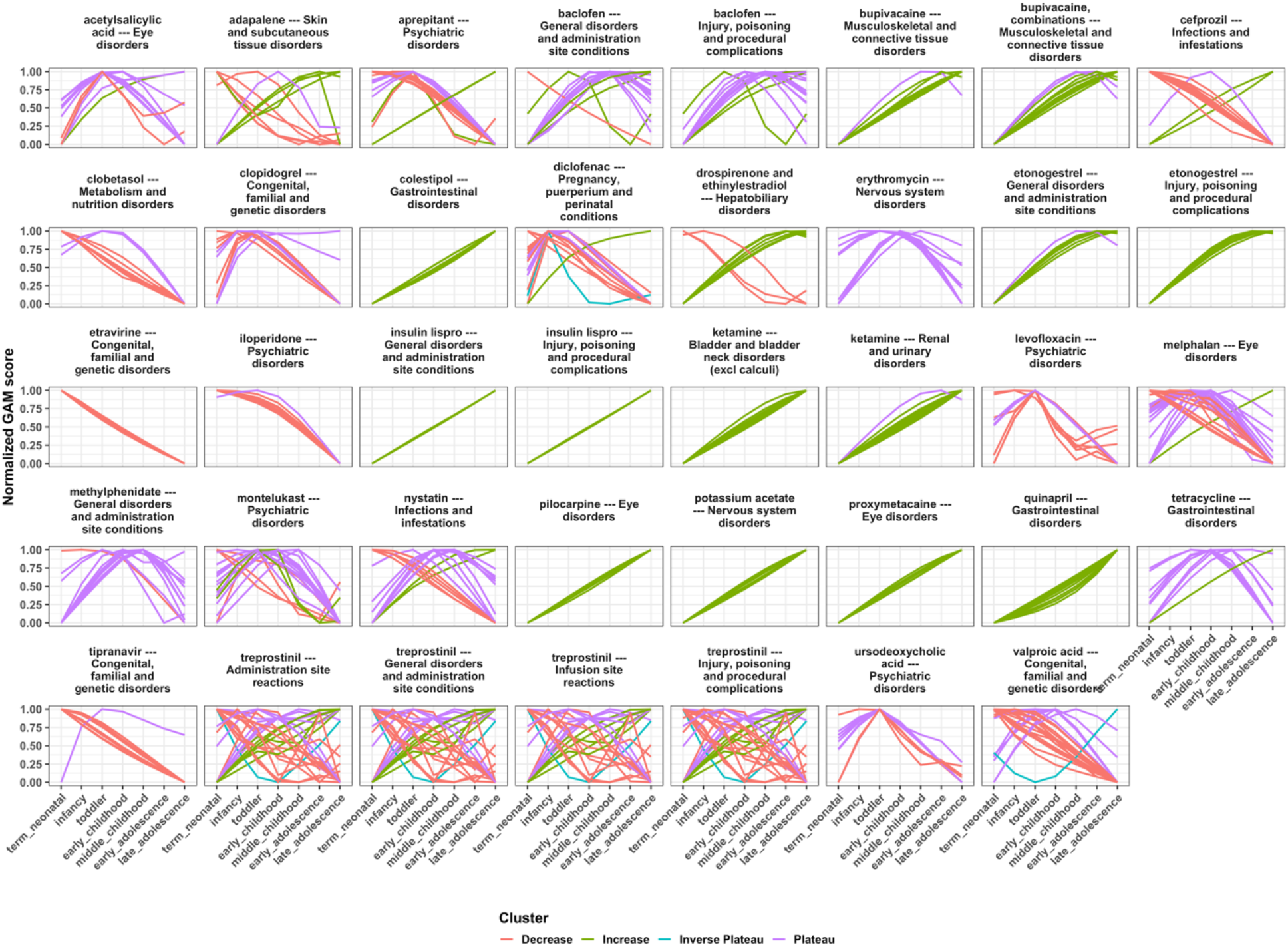
Medications posing development risks have characteristic dynamics across childhood. Drug-events were assigned risk dynamics clusters and also grouped within MedDRA system organ classes or systemic disorder. Related to Figures 4 and 5.

**Figure S5:**
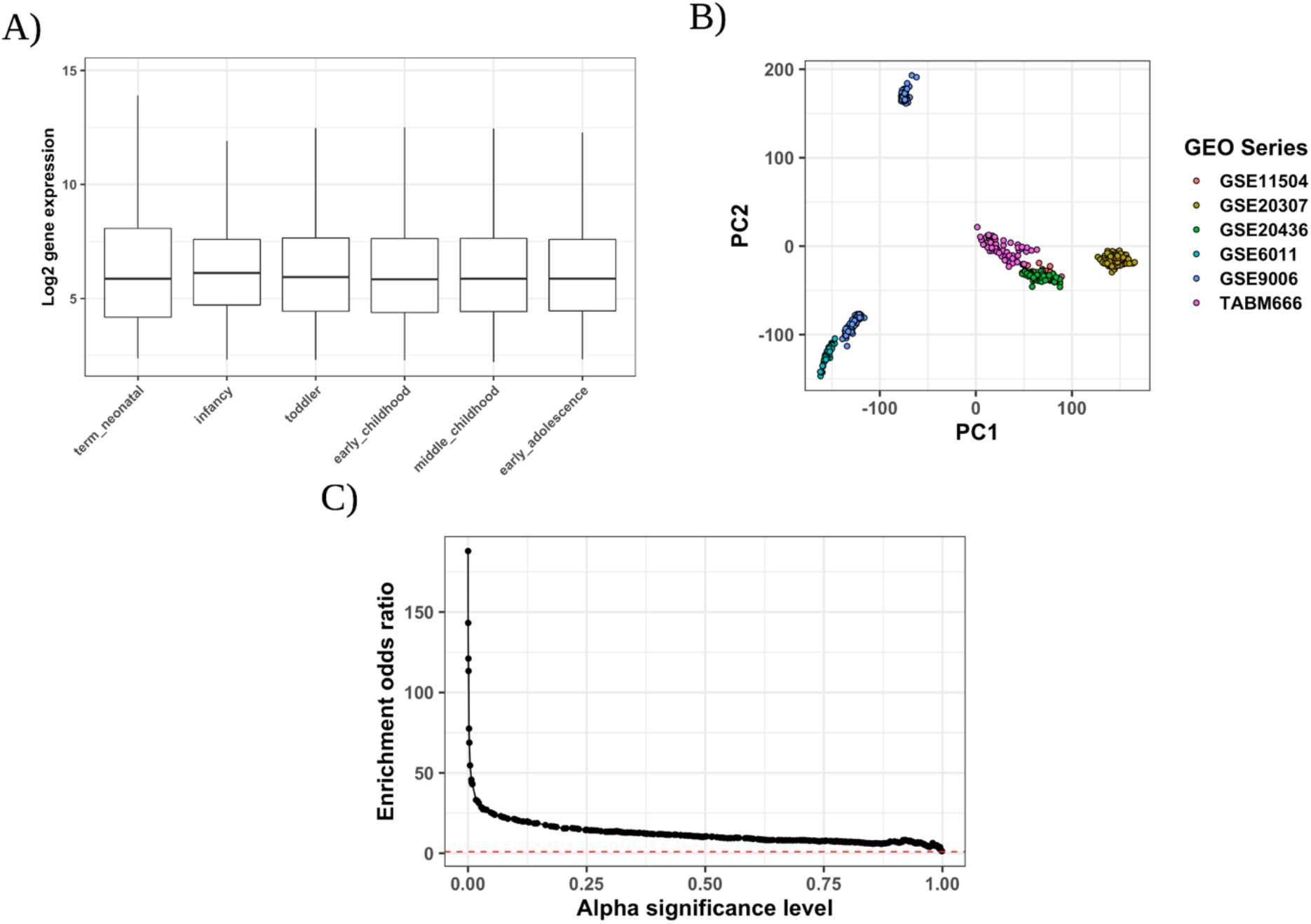
Mitigating bias in gene expression across child development stages shows association with previously found age-associated gene markers. A) Log2 probe-level gene expression across child development stages. Outliers are not shown. B) Dimension reduction of microarray expression across 576 samples and 39,558 probes. Dimensions were included in deriving stage-association. C) Enrichment analysis, at varying alpha significance thresholds, from our final gene expression dataset for the 690 significantly age-associated genes from the original Stevens et al. study. All alpha levels from the range of p-values, in tenth increments, are shown. Related to Figure 6.

**Figure S6:**
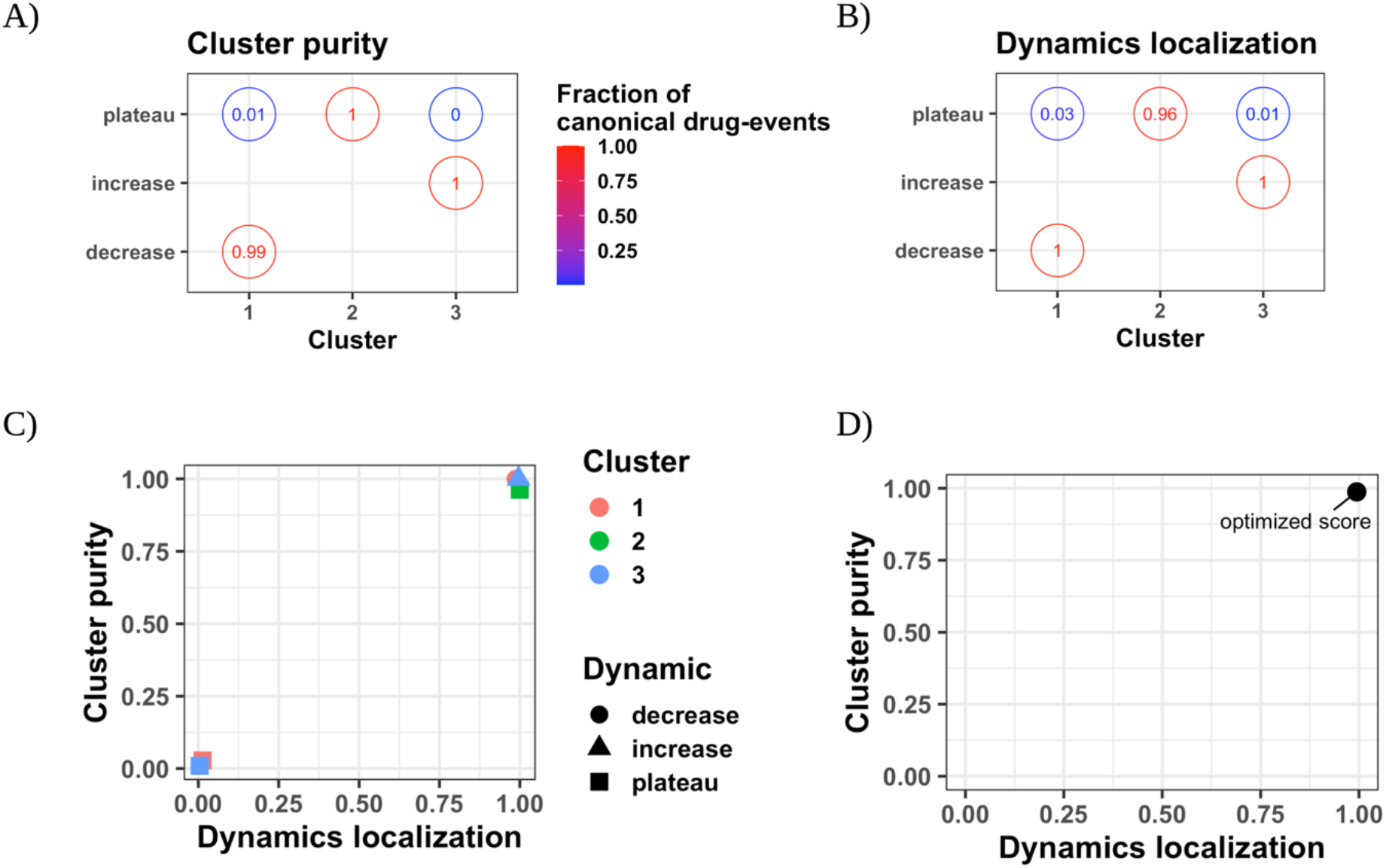
Clustering performance captures purity and localization of canonical dynamic drug-events in clusters. A) The cluster purity confusion matrix where each value is the fraction of canonical drug-events assigned to each cluster out of all drug-events in each cluster. Example breakdown for a hyperparameter set including the number of clusters K=3. The numbers shown are rounded to the second significant digit. B) The dynamics localization confusion matrix where each value is the fraction of canonical drug-events assigned to each cluster out of all drug-events in each canonical dynamics’ category. C) The product of the maximum cluster purity and dynamics localization score for each (cluster, canonical dynamics) pair. D) The overall cluster purity and dynamics localization score for a hyperparameter set. The overall score is derived in a two-step calculation from the scores in A and B): We identify the maximum cluster purity for each (cluster, canonical dynamics) pair and then average the cluster purity across canonical dynamics categories and the dynamics localization across clusters, respectively. This ensures the optimized score for each hyperparameter set scores homogeniety in both the canonical dynamics cluster assignment and the type of drug-events contained in clusters. This performance strategy is agnostic to the number of clusters K in each hyperparameter set. Related to Figure 4.

## Tables

**Table S1:**
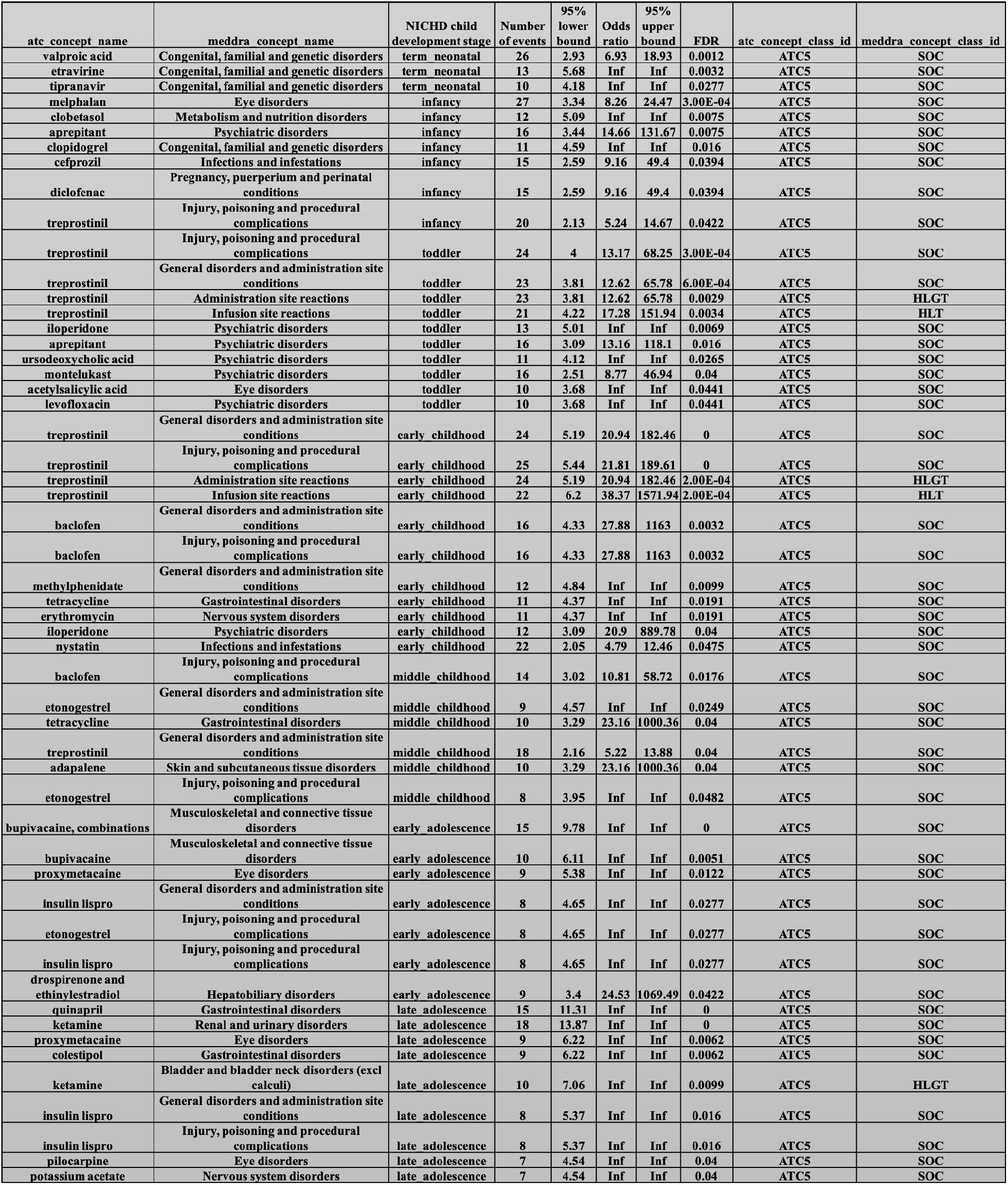
Medications pose risks for systemic disorders at child development stages. Enrichment results for the ATC 5^th^ level drugs enriched for MedDRA SOC-level events at a child development stage. The columns are ordered by child development stage then FDR. Related to Figure 4. Abbreviations: ATC5: ATC 5^th^ level; SOC: System Organ Class, HLGT: Higher-Level Group Term.

**Table S2:** The dataset of gene expression across childhood is made up of various datasets across development stages and tissues. All referenced Affymetrix image data from Stevens et al. Abbreviations: PBMCs: Peripheral blood mononuclear cells. Related to Figure 6.

| GEO series |  |  | NICHD child development stage |  |  |  |  |  |
| --- | --- | --- | --- | --- | --- | --- | --- | --- |
| GEO accession | Cell Type | Assay | term neonatal | infancy | toddler | early childhood | middle childhood | early adolescence |
| GSE11504 | Bone Marrow | hgu133plus2 | 0 | 3 | 2 | 4 | 8 | 3 |
| GSE20307 | PBMCs | hgu133plus2 | 3 | 10 | 18 | 31 | 44 | 54 |
| GSE20436 | Conjunctive Epithelium | hgu133plus2 | 0 | 4 | 5 | 20 | 29 | 2 |
| GSE6011 | Muscle | hgu133a | 0 | 22 | 7 | 6 | 2 | 0 |
| GSE9006 | PBMCs | hgu133a/b | 0 | 0 | 8 | 18 | 100 | 108 |
| TABM666 | PBMCs | hgu133plus2 | 0 | 0 | 4 | 42 | 14 | 0 |

## Availability of Data and Materials

The data used in this study is publically available from the openFDA platform. The code for this study is available https://github.com/ngiangre/pediatric_ade_database_study. The database generated by this study, in a redacted version due to size constraints, can be found at https://github.com/ngiangre/pediatric_ade_database. Please contact the authors for full data access.

## STAR Methods

### Pediatric FAERS

We created “Pediatric FAERS” as a subset of the existing FAERS database limited to those patients aged 21 or younger. We retrieved drug event reports from the Food and Drug Administration’s openFDA^43^ download page, utilizing an API key with extended permissions, containing the FAERS data. The data is comprised of safety reports listing at least one drug and at least one adverse event. Using custom python notebooks and scripts available in the ‘openFDA_drug_event-parsing’ github repository (DOI: 10.5281/zenodo.4464544), we extracted and formatted all drug event reports prior to the third quarter of 2019. Data fields included the safety report identifier, age value, age code e.g. year, adverse event the Medical Dictionary of Regulatory Activities concept code (preferred terms), and drug RxNorm code (various) used in our analyses. The age value was standardized to year units for categorizing reports into the 7 child development stages according to the Eunice Kennedy Shriver National Institute of Child and Human Development^44^. Adverse drug event MedDRA codes were mapped to standard concept identifiers using concept tables^45^ from the OMOP common data model. The drug RxNorm code was similarly translated to the standard RxNorm concept identifier (ingredient level) in OMOP and was further mapped to the equivalent Anatomical Therapeutic Chemical (ATC) Classification concept identifier (ATC 5^th^ level) using the concept relationship table. The occurrence of an adverse drug event is defined as any safety report where both the adverse event and drug concepts are reported together. The pediatric report space for any adverse drug event is all reports which have age above zero and less than or equal to 21 years old which is the upper bound for the late adolescence child development stage. All pediatric reports reported either Female or Male sex, contained the type of reporter (Physician, Consumer, lawyer, or other health professional), and the date of the report. Additionally, we joined the higher-level ATC class for drugs from the drugbank database^46^, with code to generate the database on GitHub (DOI: 10.5281/zenodo.4464604).

### ADE detection models

We compared two different models for detecting adverse drug events from spontaneous reports. First, we applied the logistic generalized additive model^47^ (GAM) to all unique drug-event pairs in Pediatric FAERS. The drug-event GAM was used to quantify adverse event risk due to drug exposure versus no exposure across child development stages. We refer to the ‘base’ GAM formula as:

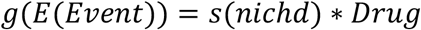

where *g* is a logit link function, *E*(*Event*) is the expected value of event reporting, *s* is a spline function with a penalized cubic basis, *nichd* is the child development stage of the report’s subject, and *Drug* is an indicator i.e. 0 or 1 of drug reporting. Details for GAMs can be found at references^48, 49^ and we specified the model using the *mgcv* package in R. Furthermore, we evaluated different model types or GAMs including different combinations of covariates: the smooth interaction effect between the report’s subject being in a child development stage and reporting female or male sex (‘Sex’), the date of first reporting the drug-event (‘Date’), the type of reporter for the drug-event (‘Reporter’), the number of drugs in ATC level 1 pharmacological drug classes for the report’s subject (‘ATC’), the number of drugs in ATC level 2 therapeutic drug classes for the report’s subject (‘ATC3’), the exposure of a drug within ATC level 1 pharmacological drug classes for the report’s subject (‘ATCbin’), the exposure of a drug within ATC level 2 therapeutic drug classes for the report’s subject (‘ATC3bin’), and a smooth effect for the number of drugs taken by the report’s subject (‘NdrugsS’).

Briefly, the GAM is a flexible statistical model that captures nonlinear effects of covariates onto a response. In this paper, we model the effect by the child development stage interacting with drug reporting on the reporting of an event where the event is the reporting of the MedDRA preferred term and the drug is the reporting of the ATC 5^th^ level drug concept. The *s*() function is a spline function where the interaction of the child development stage (main effect) and the drug (interaction using the ‘by’ variable) is modeled according to a set of basis functions. Each development stage defines the knot (7 in total) in which the expectation of event reporting is quantified. In the spline function, a penalized cubic spline basis (bs=’cs’) is used for fitting the basis functions where the first and second derivative of the event expectation is zero at each knot, resulting in a smooth event expectation across stages. To mitigate overfitting or ‘wiggliness’, we used a penalized iterative restricted likelihood approach, called ‘fREML’, with a wiggliness penalty in the objective function. Fitting the GAM model (using the ‘bam’ function and discrete=T) produces coefficient terms, similar to beta coefficients in logistic regression, for each child development stage for the association of the adverse event being reported in interaction with reporting the drug. We generated GAM scores for each child development stage resulting in 7 scores for each drug-event pair. It is important to note that all GAM scores produced were finite, nonzero values.

In addition, we made a comparison to the Proportional Reporting Ratio (PRR):

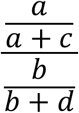

where ‘a’ is the number of reports with the drug and event, ‘b’ is the number of reports without the drug and with the event, ‘c’ is the number of reports with the drug and without the event, and ‘d’ is the number of reports without the drug or event of interest. The resulting score is the event reporting prevalence with the drug compared to without the drug. We generated PRR scores for each child development stage resulting in 7 scores for each drug-event pair.

We determined the lower confidence bound in which the population-based score would be greater than 90% of score replicates. A risk was nominally or statistically significant if the score had a 90% lower bound above the null association (null association: GAM==0, PRR==1). The drug-events from Pediatric FAERS were nominally significant if at least one risk, the risk coefficient’s 90% lower bound, was above the null association. The GAM coefficients and PRR scores were normalized between [0,1] producing scores across childhood for each drug-event to generate normalized scores.

We derived a null GAM to evaluate significance of drug-event risk compared to random drug and event reporting. We randomized drugs and events for reports, maintaining the report characteristics, and then recalculated the drug-event GAMs for 10,000 randomly selected drug and event pairs. The null GAM risk coefficients for each stage resulted in a null distribution of risks for randomly-associated drugs and events. A risk was significant by the null model if the score had a 90% lower bound above the 99% percentile of the null GAM coefficient distribution at a stage. The drug-events from Pediatric FAERS were significant by the null model if at least one risk, the risk coefficient’s 90% lower bound, was above the 99% of the null distribution for that stage. This ensured that at least one risk at a child development stage was nominally significant as well.

Unless otherwise specified, all statistics in brackets are the lower and upper 90% confidence intervals.

### ADE detection model likelihood and performance

We evaluated the generalizability and model likelihood i.e. fit of a drug-event GAM, including each model type, on a random sample of 2,000 drug-events from Pediatric FAERS. We quantified model statistics using a proportion of the 2,000 drug-events that were reported 50 or more times and an additional, complementary set of drug-events with less than 50 reports. We quantified the fit of the drug-event GAMs using the Akaike’s Information Criterion (AIC), which is a measure of the tradeoff between model likelihood and complexity. We quantified the generalization of the drug-event GAMs by fitting each model on 80% of the dataset, termed the training set, and quantifying the area under the receiver operating characteristic (AUROC) curve on both the training set and the unseen testing set. The training and testing sets were balanced in having the same proportion of reports with (20%) and without (80%) the adverse event. Each model type was fit on the same training set.

### Drug-event reference sets

#### Ryan et al. reference set

We downloaded the adverse event reference set developed by Ryan et al.^50^ containing manually curated positive and negative control drugs associated to four outcomes: acute liver injury, acute kidney injury, acute myocardial infarction, and upper gastrointestinal bleeding. We mapped RxNorm concepts to ATC 5^th^ level. There were 1078, 59, 284, and 928 unique adverse drug events, both positive and negative controls, for each outcome, respectively. We found 312, comprising only three outcomes, out of 2,349 total drug-event pairs (13%) within Pediatric FAERS. There were 74 negative and 238 positive drug-event pairs.

#### GRiP pediatric reference set

We extracted the drug-event pairs observed in Pediatric FAERS listed within the pediatric drug-event reference standard from the Global Research in Pediatrics consortium^31^. A machine-readable dataset can be found at the ‘GRiP_pediatric_ADE-reference_set’ github repository (DOI: 10.5281/zenodo.4453379). We assigned drug-event pairs with epidemiological or mechanistic evidence in children (Control==’C’ and Control==’B’) as the positive class (N=179 and 75 in Pediatric FAERS), and the cross-product of all drugs and events that were complementary to drug-event pairs in the reference set as the negative class (N=397 and 112 in Pediatric FAERS). In total, we evaluated 187 positive and negative drug-events observed in Pediatric FAERS.

#### Pediatric adverse events

We downloaded and joined standard concepts to the pediatric adverse event term list from the MedDRA website (https://www.meddra.org/paediatric-and-gender-adverse-event-term-lists). This term list is no longer supported but we provide a machine-readable version in our resource.

### ADE clustering

We identified clusters of risk patterns across development stages for all drug-events in Pediatric FAERS. We considered ADE risk patterns as time series, representing temporal drug-event risk across child development stages. The temporal risk scores are the normalized, between 0 and 1, GAM scores of the interaction between child development stage and drug exposure. We used the R package *dtwclust*^51^ to compare different distance and centroid methods due to ease of implementation and optimization of computationally expensive methods. We performed an iterative procedure to evaluate the clustering by different combinations of distance and centroid methods.

We used a partitional clustering strategy, which minimizes the intra-cluster distance while maximizing the inter-cluster distance by iterative greedy descent to converge to a local optima^52^. The distance methods evaluated were dynamic time warping (DTW), which is a fast implementation to find the optimum warping path between two drug-events (‘dtw_basic’); the shape-based distance (‘sbd’), which is a shift and scale-invariant comparison of time series based on the k-Shape algorithm^53^; and the triangular global alignment kernel (‘gak’) which is a kernel method unlike DTW that has been shown faster and more efficient in classification tasks^54^. The centroid methods that evaluated cluster assignment for drug-events were the average risks between drug-events across childhood (‘mean’); the partition-around-medoids (‘pam’), which utilizes one of the series as the cluster centroid; DTW barycenter averaging (‘dba’), which finds the optimum average drug-event between drug-event series in DTW space; and shape averaging (‘shape’) based on the k-Shape algorithm, which extracts the most representative drug-event dynamic to utilize as the centroid^53^.

We fit the clustering model, with hyperparameter sets that include a distance metric, centroid method, and number of clusters K, on a random sample of 10,000 drug-events from Pediatric FAERS along with spiked-in ‘canonical’ dynamics patterns. We assert that reporting dynamics during childhood reflect ontogenic profiles observed on molecular, functional, and structural levels^4, 55, 56^. The canonical drug-events previously studied, including filtering for the ranked pattern of interest, were categorized as ‘increase’ (N=237), ‘decrease’ (N=224) , or ‘plateau’ dynamics (N=106)^1^. We quantified clustering performance for each hyperparameter set using the drug-events in each canonical dynamics’ category and their cluster assignment (see Figure S5 for details and illustrations of the strategy). Overall, we developed two custom metrics: 1) Cluster purity, which is the clustering precision or the score for drug-events from a canonical dynamics category assigned to a cluster, and 2) Dynamics localization, which is the clustering sensitivity or the score for drug-events within a cluster from a particular canonical dynamics’ category. This ensured the clustering performance , both the cluster purity and dynamics localization, for each hyperparameter set scores the homogeniety in both the assignment of dynamics and type of dynamics within clusters. We only considered the predominant or most frequent cluster of each of the three dynamics to compute the above metrics, allowing for comparing performance for K>3 (see Figure S6 for details and illustrations of the strategy).

### ADE cluster enrichment

We evaluated the enrichment of drug-events, which were significant by the null model, within clusters and categories of drug-events. We calculated the fisher’s exact test to evaluate enrichment of drug-events within a specific category to also be assigned a specific cluster.

### ADE stage enrichment

We evaluated the enrichment of drug-events, which were significant by the null model, within child development stages and categories of drug-events. We calculated the fisher’s exact test to evaluate enrichment of drug-events within a specific category to also have a significant risk, by the null model, at a specific stage.

### Pediatric gene expression dataset

#### Dataset processing

We extracted expression data from GEO and EBI microarray datasets utilized by Stevens et al.^8^ to derive gene expression across child development stages (Table S2). We compiled datasets’ raw image (CEL) files (affymetrix images only) and integrated annotation to the microarray probe sets. We used the Bioconductor R package *affy* to load the microarray data.

Samples were preprocessed together per the same assay (hgu133a, hgu133b, and hgu133plus2) closely aligning to the procedure in Stevens et al. Specifically, we used Robust Microchip Average (RMA) background correction (‘rma’), quantile normalization (‘quantile’), perfect match correction algorithm (‘pmonly’), and mean probe set summarization (‘avgdiff’). We used the R package *ROMOPOmics* (DOI: 10.5281/zenodo.4463257) to extract phenotypic data from each GEO dataset (the ‘TABM666’ dataset was downloaded from the EBI website) and convert the age of each sample from the datasets to year units. We defined the NICHD child development stages using the age of the samples within the stage age boundaries^44^. We mapped probe set IDs to uniprot IDs and to gene symbols using the libraries of each assay’s annotation R package within Bioconductor. We evaluated the average difference in log2 probe expression values, using 10,000 samples with replacement, by Students t-test between each adjacent child development stage.

#### Dataset validation

We performed a validation analysis to evaluate the processed gene expression data to reproduce the significant age-associated findings published by Stevens et al^8^. Specifically, there were 690 genes comprising 927 probes with expression that was significantly associated to age. We then performed an association analysis to evaluate significant stage-associated genes from our gene expression dataset (we only utilized the datasets from their main analysis dataset: GSE11504, GSE9006, and TABM666). We quantified stage-association of each probe’s expression using a GLM where the probe value was the dependent variable and the NICHD stage, where each category was an integer, and the first six principal components and GEO series indicator minus the intercept term were predictors or covariates. We computed the odds ratio using the hypergeometric test to compare overlap of age-associated and stage-associated genes, compared to those that were not, at different alpha significance thresholds (we required at least one probe to be significantly stage-associated per gene). We found that the enrichment of genes in our data to be robust at varying significance levels (Figure S4C). The robust enrichment provided evidence that our gene expression dataset was capturing accurate dynamic expression patterns across childhood.

### Pediatric cytochrome P450 gene expression dynamics evaluation

We evaluated whether drug-event risk dynamics were dependent on expression dynamics of the drug’s substrate using our pediatric gene expression dataset. Again, we only utilized the datasets from their main analysis dataset: GSE11504, GSE9006, and TABM666.

First, to account for observed batch effects (Figure S4B), we performed a regression for every probe to mitigate bias of gene expression dynamics across child development stages and all datasets. We used the residual probe levels from a GLM where the dependent variable were the observed values, the observations were each sample, and the covariates were the first six principal components and the GSE series indicator. Importantly, the GLM did not include association to development stages so that the residuals capture the difference between observed probe values and the batch-predicted probe values (again, we only utilized the datasets from their main analysis dataset: GSE11504, GSE9006, and TABM666). We used these probe residual values in our downstream analysis.

We identified cytochrome P450 gene products using a regex expression ‘^CYP’ on gene symbols to extract probe-level expression data. We performed a correlation analysis between pairs of probes within non-random *CYP* gene products and removed probes that showed a negative correlation (Pearson r<0) in at least one pairwise comparison.

We manually scraped drugbank webpages to determine the mapping between drug enzymes and uniprot IDs. We then filtered for drugs that were annotated as substrates for *CYP* gene products, again using the gene symbol pattern matching to the regex expression ‘^CYP’. We only considered drug risks where side effects were listed on the drug’s label according to SIDER 4.0^57^ (N=780).

We hypothesized expression of *CYP* enzymes across child development stages influence the adverse event risks of drugs they metabolize versus do not metabolize. In other words, drugs that were substrates for *CYP* enzymes generated drug-event risks with more shared information with expression dynamics than drugs tthat were not substrates. We generated the two distributions by computing the mutual information (MI), using the *maigesPack* R package in Bioconductor, between the *CYP* probes’ residual expression, averaged across samples, and drug risks’ score for each child development stage, if both expression and risk were present. We generated a distribution of mutual informations from (drug-event, *CYP* gene probe, z score) triplets. The drug-event risk score at each stage varied according to a randomly selected z-score from a standard normal distribution. We used the mean (mu) and the standard error (SE) of the GAM estimates to generate a new risk score at each child development stage:

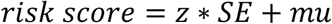

These permutations generated substrate MIs and non-substrate MIs for each *CYP* enzyme. The two MI distributions were compared using Student’s t-test to evaluate a greater difference in average mutual information for substrate compared to non-substrate drug-event risks for their substrate’s gene expression. Also, we computed a Mann Whitney one-sided test to evaluate whether the substrate risk comparisons were greater in rank than non-substrate risks, on average across possible risk variations for a drug-event and *CYP* probe residual expression. We derived an AUROC statistic by normalizing the Mann Whitney U statistic for the number of comparisons between substrate (*n*_0_) and non-substrate (*n*_1_) doublets:

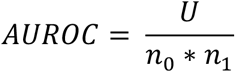

We made these comparisons across all event disorders, detailed above, as well systemic events of a system organ class by the same procedure.

